# COVID-19 mRNA Vaccination Reduces Guillain-Barré Syndrome Risk: Evidence from a Large Longitudinal Cohort Study

**DOI:** 10.64898/2026.07.02.26357163

**Authors:** Jinju Li, Yuanyi Pan, Yun Han, Chuan Zhou, Lili Zhao, Yongqun He, N3C Consortium

## Abstract

The association between COVID-19 vaccination and Guillain-Barré syndrome (GBS) has been previously investigated with inconsistent results, largely due to limited data and lack of concurrent controls. To address this problem, a large longitudinal cohort study was conducted using National COVID Cohort Collaborative (N3C) data. While COVID-19 infection was associated with increased GBS occurrence, COVID-19 vaccination was associated with significantly reduced GBS risk relative to unexposed (unvaccinated and uninfected) control, corresponding to a 61% lower 30-day risk (incidence risk ratio: IRR = 0.39, *P* < 0.01), consistent with multivariable Cox regression showing a similar reduction (adjusted hazard ratio: aHR = 0.41, *P* < 0.01). This protective association was observed only among recipients of mRNA vaccines (BNT162b2: IRR = 0.38, *P* < 0.01; mRNA-1273: IRR = 0.24, *P* < 0.01), but not among recipients of adenoviral-vector vaccines (IRR = 1.38, *P* > 0.05). Prior COVID-19 vaccination also reduced infection-associated GBS risk. Additional factors associated with GBS risk included sex, vaccine dose, and pre-existing comorbidities such as stroke, neurological disorders, and autoimmune diseases. Overall, our N3C large-scale study provides evidence that COVID-19 mRNA vaccination reduces GBS risk, supporting the safety profile of mRNA vaccines and warranting further mechanistic investigation.

## Introduction

The emergence of SARS-CoV-2 and the ensuing COVID-19 pandemic triggered a global public health crisis, leading the World Health Organization to declare a Public Health Emergency of International Concern on January 30, 2020.^1^ The urgent need to control the pandemic accelerated the development and deployment of COVID-19 vaccines, including novel mRNA and adenoviral vector platforms, which demonstrated high efficacy and safety in large randomized clinical trials.^2,3^ However, rare adverse events may not be captured in these trials, making robust post-marketing surveillance essential for evaluating real-world vaccine safety.^4,5^

Amid the global COVID-19 vaccination campaign, concerns arose about rare but serious neurological complications, including Guillain–Barré syndrome (GBS),^6^ a potentially life-threatening polyradiculoneuropathy characterized by rapidly progressive weakness and sensory deficits. While GBS is rare, it can be life-threatening and debilitating and has historically been associated with both infections and certain vaccines.^7–9^ GBS incidence increased during the pandemic, with COVID-19 associated cases reported to be more severe than non-COVID-related cases.^10–13^ Case reports have also documented GBS following both mRNA and adenoviral vector COVID-19 vaccinations in the early stage of pandemic. ^14–18^ However, evidence regarding the relationship between COVID-19 vaccination and GBS remains inconsistent. Pharmacovigilance data from surveillance systems such as the Vaccine Adverse Event Reporting System (VEARS) or VigiBase have identified a potential increase in GBS risk following adenoviral vector vaccines, including Ad26.COV2.S and ChAdOx1 nCoV-19, studies evaluating mRNA vaccines have generally found minimal or no excess risk as they compared the incidence of GBS associated with mRNA vs. adenoviral-vector vaccination.^19–24^ But VAERS is a passive, self-reported adverse event surveillance system that lacks a well-defined denominator, limiting its ability to estimate incidence rates or establish causal associations. It has been suggested that GBS incidence following the Pfizer-BioNTech mRNA vaccine was consistent with expected background rates; however, this finding should be interpreted cautiously because the background incidence of GBS among unvaccinated individuals during the COVID-19 pandemic had not yet been well characterized at the time of the study was conducted.^25^ Furthermore, a nested case control study involving 76 patients with GBS and 760 matched controls has suggested that mRNA vaccines may confer a protective effect against GBS, potentially through reducing the risk or severity of COVID-19 infection.^26^ These divergent findings have contributed to ongoing uncertainty regarding COVID-19 vaccines associations with GBS.

The inconsistent results are likely due to multiple methodology limitations. Due to limited available clinical data during the early phase of the pandemic for emergence of a novel virus, previous studies on COVID-19 vaccination associated GBS risk primarily relied on comparisons between vaccine platforms and did not account for differences in vaccine effectiveness in preventing COVID-19 infection.^22–24^ This is an important limitation, as differences in vaccine efficacy may introduce confounding by competing risk, as recipients of lower-efficacy vaccines are more likely to experience breakthrough COVID-19 infection, and the failure to account for participates with infection in these studies may bias the results, given that infection itself is a trigger for GBS.^2–4, 10–13^ Additionally, many studies relied on comparisons with historical background rates rather than contemporaneous unvaccinated control groups and did not account for temporal differences in SARS-CoV-2 circulation or prior vaccination of COVID-19 infection.^22–24^ A further limitation is that most studies examined only narrow risk windows like 21, 42 days rather than a longitudinal time-to-event framework to fully capture the associations.^21–28^

To address these gaps, we utilized data from the National Clinical Cohort Collaborative (N3C), one of the largest and most representative longitudinal electronic health record (EHR) databases in the United States, covering the period from December 1, 2020, to November 1, 2024.^29,30^ To better isolate vaccine-attributable risk, we excluded GBS cases preceded by COVID-19 infection and those occurring within the follow-up windows and applied a time-to-event framework with concurrent unvaccinated control. Our objectives were to characterize the incidence and clinical profile of post-vaccination GBS, compare risk across vaccine platforms, and evaluate whether prior vaccination modifies GBS risk following subsequent COVID-19 infection, thereby generating evidence-based insights to inform vaccine safety surveillance and public health decision-making.

## Results

### Study Population

The study included 8,474,761 adults, comprising 3,880,554 individuals in the COVID-19 vaccination group, 4,217,359 in the COVID-19 infection group, and 376,848 in the control group (Table 1). Among vaccinated individuals, 83.03% received mRNA vaccines, 5.03% received adenovector vaccines, and 11.94% received other vaccine types. A total of 56 GBS cases were identified in the vaccination group (incidence rate: 1.44 per 100,000 persons; 95% CI: 1.06–1.82), compared with 336 cases in the COVID-19 infection group (incidence rate: 7.97 per 100,000 persons; 95% CI: 7.13–8.87) and 14 cases in the control group (incidence rate: 3.71 per 100,000 persons; 95% CI: 2.02–6.14) (see Figure 1).

**Figure 1:**
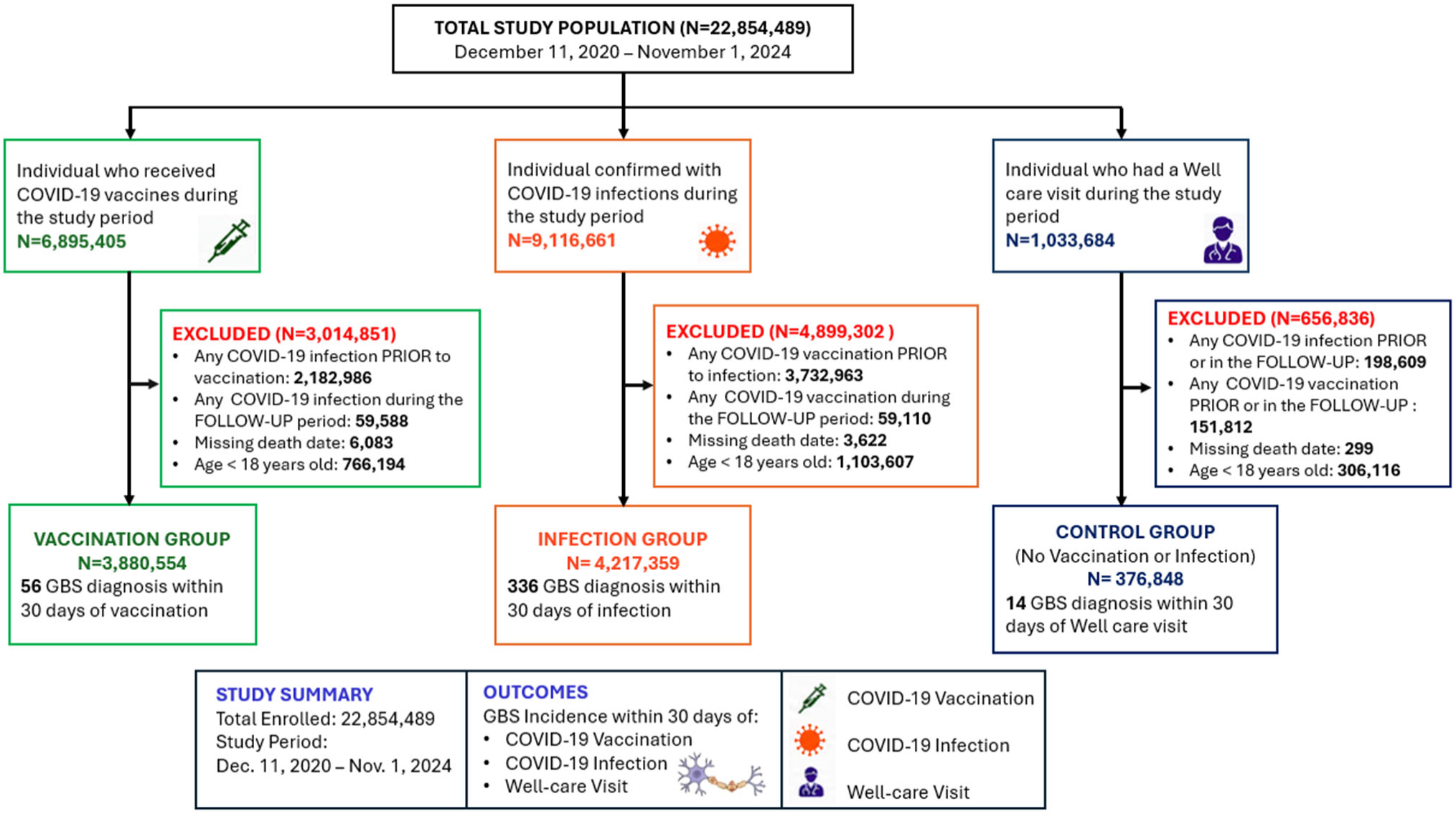
Flowchart of Development of Analysis Cohorts Based on Initial COVID-19 Antigen Exposure. The study participants were categorized into three groups based on their initial exposure to COVID-19 antigens: the vaccination group, the infection group, and the control group without any vaccination or infection during the period from December 11, 2020 to November 1, 2024. The vaccine group: Those who received COVID-19 vaccines were included, after excluding any preceded infection or follow-up period infection; The infection group: Those who were confirmed with COVID-19 infection after December 11, 2020 were included, after excluding any preceded vaccination or follow-up period vaccination. The control group: Those who took well-care visits after December 11, 2020 were included, after excluding any preceded vaccination and infection or follow-up period vaccination and infection. Abbreviation: GBS, Guillain-Barré Syndrome.

**Table 1:**
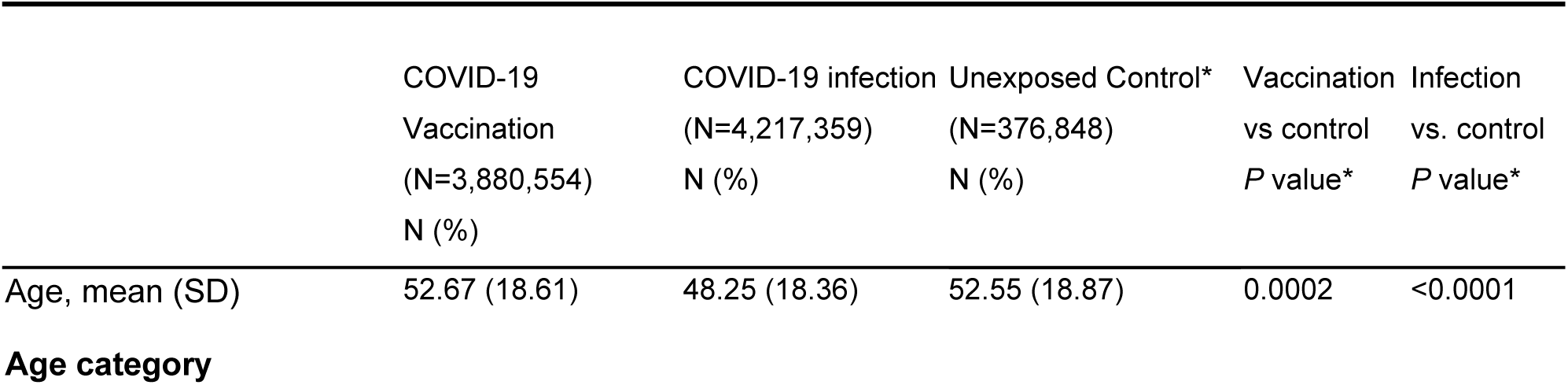
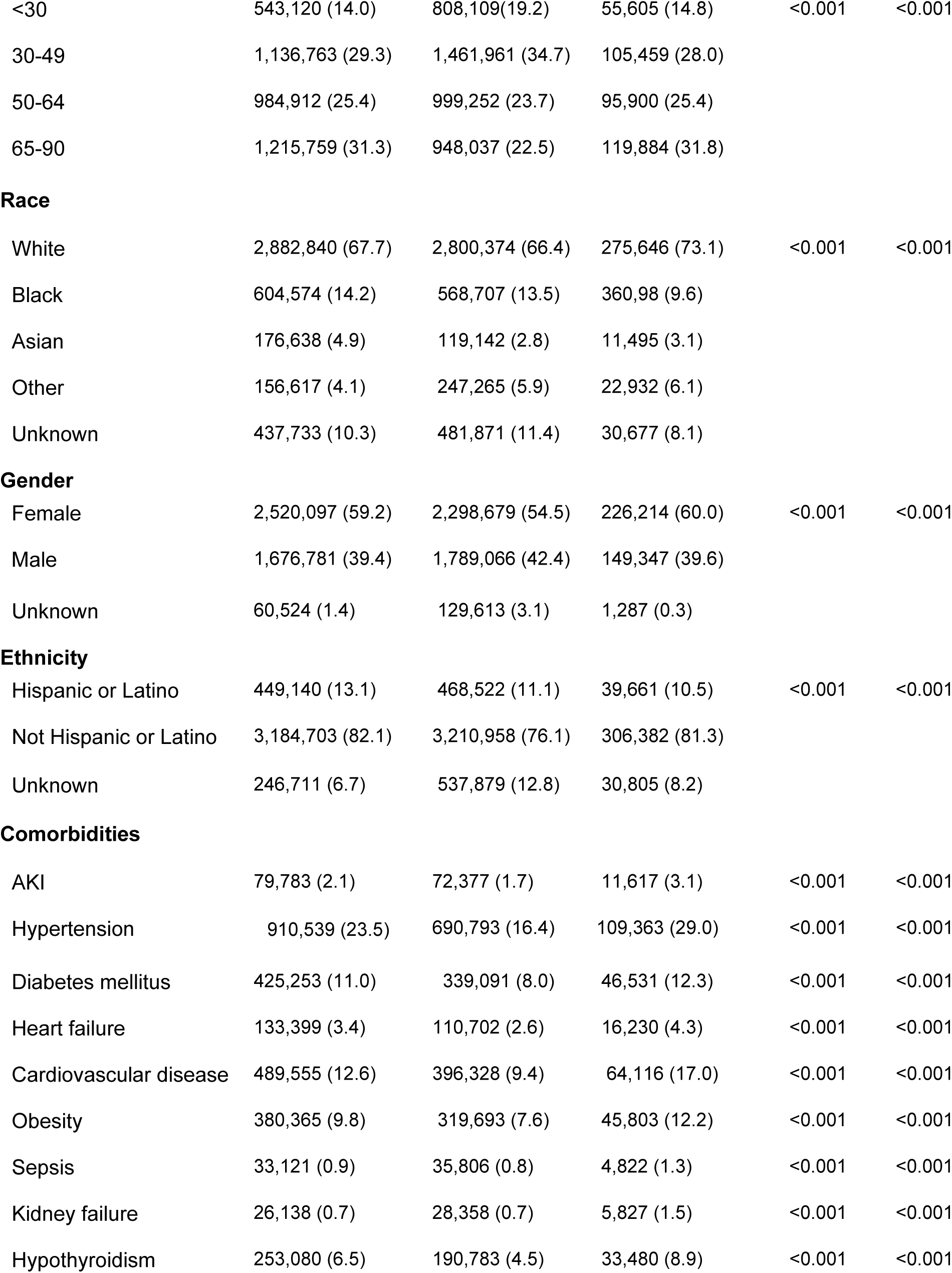
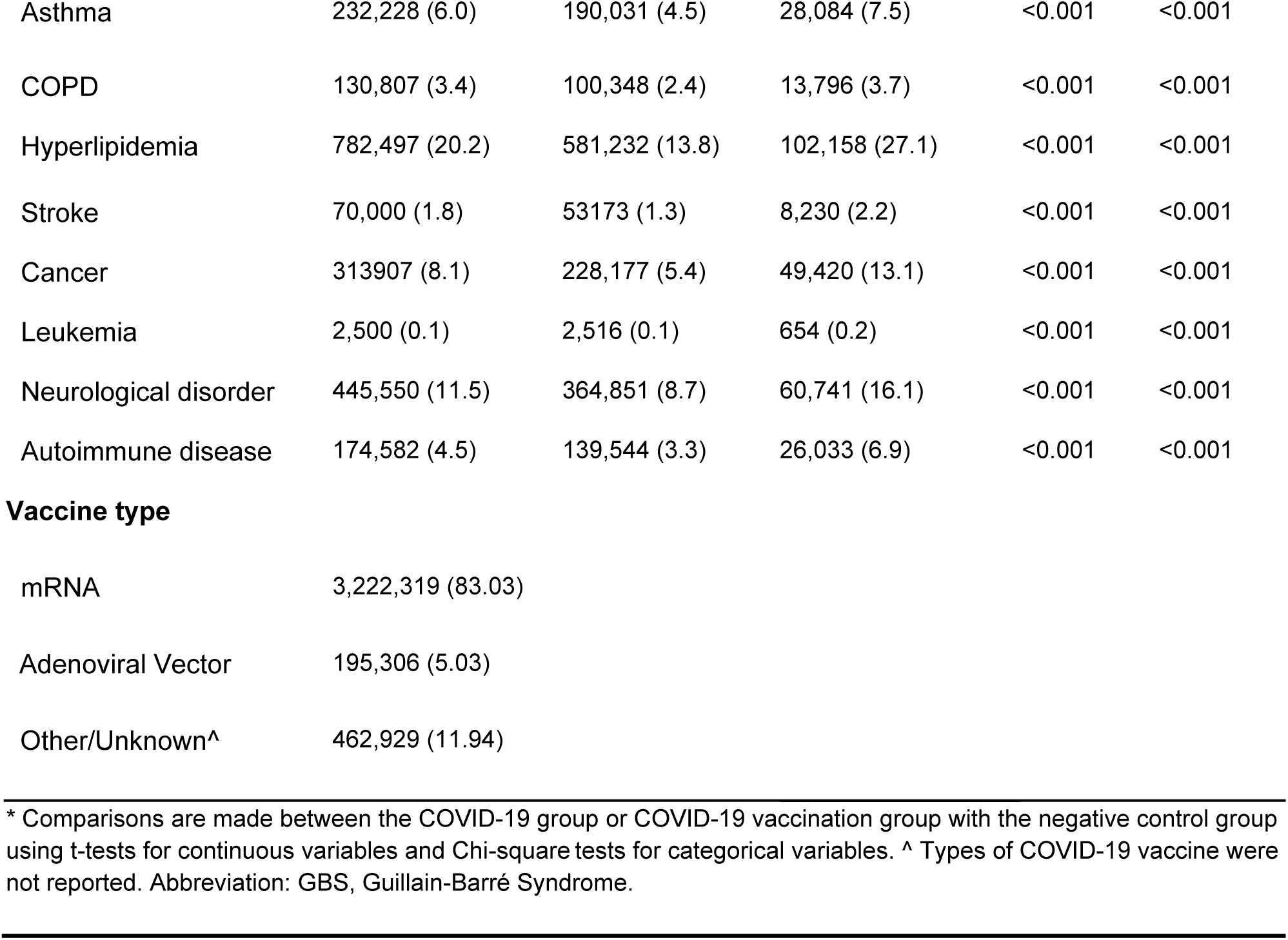
Baseline Characteristics of Patients with COVID-19 vaccination, infection and control groups from December 11, 2020 to November 1, 2024 in N3C.

|  | COVID-19<br>Vaccination<br>(N=3,880,554)<br>N (%) | COVID-19 infection<br>(N=4,217,359)<br>N (%) | Unexposed Control*<br>(N=376,848)<br>N (%) | Vaccination<br>vs control<br>*P* value* | Infection<br>vs. control<br>*P* value* |
| --- | --- | --- | --- | --- | --- |
| Age, mean (SD) | 52.67 (18.61) | 48.25 (18.36) | 52.55 (18.87) | 0.0002 | <0.0001 |
| **Age category** |  |  |  |  |  |
| <30 | 543,120 (14.0) | 808,109(19.2) | 55,605 (14.8) | <0.001 | <0.001 |
| 30-49 | 1,136,763 (29.3) | 1,461,961 (34.7) | 105,459 (28.0) |  |  |
| 50-64 | 984,912 (25.4) | 999,252 (23.7) | 95,900 (25.4) |  |  |
| 65-90 | 1,215,759 (31.3) | 948,037 (22.5) | 119,884 (31.8) |  |  |
| **Race** |  |  |  |  |  |
| White | 2,882,840 (67.7) | 2,800,374 (66.4) | 275,646 (73.1) | <0.001 | <0.001 |
| Black | 604,574 (14.2) | 568,707 (13.5) | 360,98 (9.6) |  |  |
| Asian | 176,638 (4.9) | 119,142 (2.8) | 11,495 (3.1) |  |  |
| Other | 156,617 (4.1) | 247,265 (5.9) | 22,932 (6.1) |  |  |
| Unknown | 437,733 (10.3) | 481,871 (11.4) | 30,677 (8.1) |  |  |
| **Gender** |  |  |  |  |  |
| Female | 2,520,097 (59.2) | 2,298,679 (54.5) | 226,214 (60.0) | <0.001 | <0.001 |
| Male | 1,676,781 (39.4) | 1,789,066 (42.4) | 149,347 (39.6) |  |  |
| Unknown | 60,524 (1.4) | 129,613 (3.1) | 1,287 (0.3) |  |  |
| **Ethnicity** |  |  |  |  |  |
| Hispanic or Latino | 449,140 (13.1) | 468,522 (11.1) | 39,661 (10.5) | <0.001 | <0.001 |
| Not Hispanic or Latino | 3,184,703 (82.1) | 3,210,958 (76.1) | 306,382 (81.3) |  |  |
| Unknown | 246,711 (6.7) | 537,879 (12.8) | 30,805 (8.2) |  |  |
| **Comorbidities** |  |  |  |  |  |
| AKI | 79,783 (2.1) | 72,377 (1.7) | 11,617 (3.1) | <0.001 | <0.001 |
| Hypertension | 910,539 (23.5) | 690,793 (16.4) | 109,363 (29.0) | <0.001 | <0.001 |
| Diabetes mellitus | 425,253 (11.0) | 339,091 (8.0) | 46,531 (12.3) | <0.001 | <0.001 |
| Heart failure | 133,399 (3.4) | 110,702 (2.6) | 16,230 (4.3) | <0.001 | <0.001 |
| Cardiovascular disease | 489,555 (12.6) | 396,328 (9.4) | 64,116 (17.0) | <0.001 | <0.001 |
| Obesity | 380,365 (9.8) | 319,693 (7.6) | 45,803 (12.2) | <0.001 | <0.001 |
| Sepsis | 33,121 (0.9) | 35,806 (0.8) | 4,822 (1.3) | <0.001 | <0.001 |
| Kidney failure | 26,138 (0.7) | 28,358 (0.7) | 5,827 (1.5) | <0.001 | <0.001 |
| Hypothyroidism | 253,080 (6.5) | 190,783 (4.5) | 33,480 (8.9) | <0.001 | <0.001 |
| Asthma | 232,228 (6.0) | 190,031 (4.5) | 28,084 (7.5) | <0.001 | <0.001 |
| COPD | 130,807 (3.4) | 100,348 (2.4) | 13,796 (3.7) | <0.001 | <0.001 |
| Hyperlipidemia | 782,497 (20.2) | 581,232 (13.8) | 102,158 (27.1) | <0.001 | <0.001 |
| Stroke | 70,000 (1.8) | 53,173 (1.3) | 8,230 (2.2) | <0.001 | <0.001 |
| Cancer | 313,907 (8.1) | 228,177 (5.4) | 49,420 (13.1) | <0.001 | <0.001 |
| Leukemia | 2,500 (0.1) | 2,516 (0.1) | 654 (0.2) | <0.001 | <0.001 |
| Neurological disorder | 445,550 (11.5) | 364,851 (8.7) | 60,741 (16.1) | <0.001 | <0.001 |
| Autoimmune disease | 174,582 (4.5) | 139,544 (3.3) | 26,033 (6.9) | <0.001 | <0.001 |

**Vaccine type**
|  |  |
| --- | --- |
| mRNA | 3,222,319 (83.03) |
| Adenoviral Vector | 195,306 (5.03) |
| Other/Unknown^ | 462,929 (11.94) |

\* Comparisons are made between the COVID-19 group or COVID-19 vaccination group with the negative control group using t-tests for continuous variables and Chi-square tests for categorical variables. ^ Types of COVID-19 vaccine were not reported. Abbreviation: GBS, Guillain-Barré Syndrome.

### Association Between COVID-19 Vaccination and Risk of GBS

The time-to-event analysis revealed that the risk of developing GBS was significantly higher in the COVID-19 infection group than in either the vaccination (incidence risk ratio: IRR = 6.01, 95% CI, 4.54-7.97, *P* < 0.0001, Supplementary Figure 1) or the control group (IRR = 2.34, 95% CI, 1.37-3.98, *P* = 0.004, Supplementary Figure 1). However, during the 30-day follow-up period, COVID-19 vaccination was associated with a significantly lower risk of GBS compared with the unexposed control group (IRR = 0.39; 95% CI, 0.22–0.70; *P* = 0.001; Figure 2 and Supplementary Figure 1).

**Figure 2:**
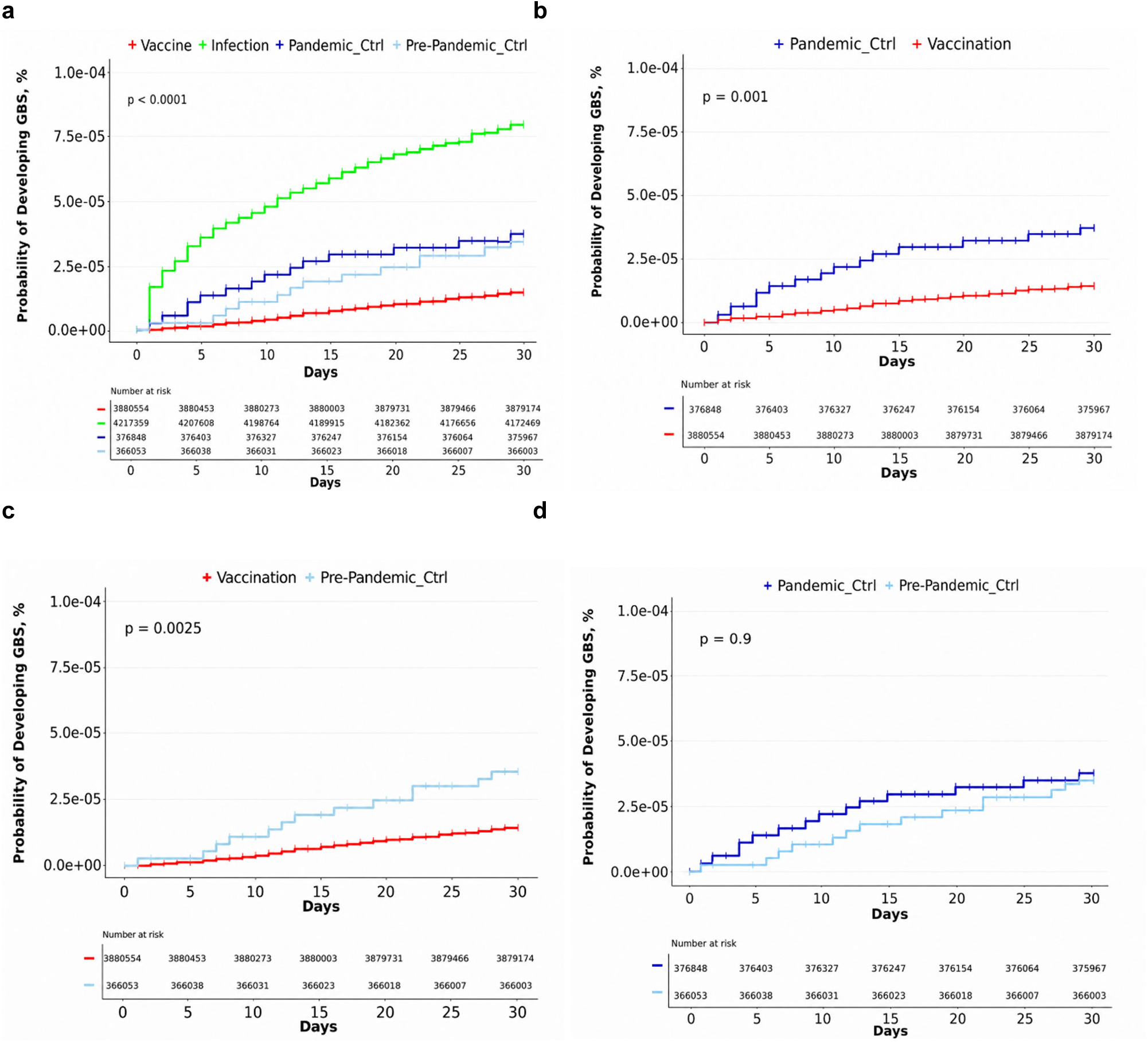
Association Between COVID-19 Vaccination and Risk of GBS Within 30 Days of Vaccination. (a) Kaplan–Meier curves representing the cumulative incidence of GBS within 30 days post-exposure across four groups: vaccination (red curve), infection (green curve), pandemic control (blue curve), and pre-pandemic control (light blue curve). (b) Cumulative curves representing the GBS diagnosis within 30 days after COVID-19 vaccination, compared with the pandemic (contemporaneous) control group without documented COVID-19 infection or vaccination in the primary analysis. (c) Cumulative curves representing GBS diagnosis within 30 days after COVID-19 vaccination compared with pre-pandemic controls (well-care visits between January 2018 and January 2020). (d) Cumulative curves representing GBS diagnosis comparing unexposed pandemic control with pre-pandemic control. The x-axis represents days from index date, and the y-axis indicates the probability of developing GBS as a percentage. Statistical significance was assessed using a Log-rank test. Abbreviation: GBS, Guillain-Barré Syndrome.

To further evaluate the robustness of the observed association and address potential healthy-user bias and residual confounding, we performed multivariable Cox proportional hazards regression analyses to estimate the association between COVID-19 vaccination and the risk of GBS after adjustment for potential confounders. Covariates included age, sex, race, ethnicity, prior healthcare utilization, and pre-existing comorbidities. This allowed us to assess the independent association between COVID-19 vaccination and GBS risk while accounting for demographic characteristics and underlying health conditions that could influence the outcome. In multivariable Cox regression analyses, COVID-19 vaccination remained independently associated with a lower risk of GBS after adjustment for potential confounders (aHR, 0.41; 95% CI, 0.23–0.75). These findings were consistent with the primary analysis and further support an association between COVID-19 vaccination and reduced GBS risk.

Given the potential for misclassification bias due to undetected COVID-19 cases within the pandemic-unexposed control group, a historical pre-pandemic control group was included as an additional comparator. The protective association between COVID-19 vaccination and reduced GBS risk remained consistent when this comparator was used: compared with the historical pre-pandemic control group, vaccinated individuals had a significantly lower risk of GBS (IRR = 0.41; 95% CI, 0.22–0.74; *P* = 0.0025; Figure 2c and Supplementary Figure 1). Notably, no significant difference in GBS incidence was observed between the pandemic and pre-pandemic controls (Figure 2d), supporting the stability of baseline GBS risk across study periods. This suggests that the observed reduction in GBS risk among vaccinated individuals is unlikely to be explained by temporal trends or background fluctuations in GBS incidence.

### Time- and Dose-Dependent Association Between COVID-19 Vaccination and Risk of GBS

Secondary analyses were conducted to assess the robustness of the findings by stratifying results according to follow-up duration and vaccine doses. Follow-up duration was examined to determine whether GBS risk differed across distinct risk windows after vaccination, while vaccine doses analyses assessed potential dose–response relationships. The protective association between COVID-19 vaccination and GBS remained statistically significant within 60 days (aHR = 0.56; 95% CI, 0.34–0.93; *P* = 0.0244). However, by 90 days, the association was no longer statistically significant (aHR = 0.66; 95% CI, 0.42–1.04; *P* = 0.0743), suggesting a time dependent effect of COVID-19 vaccination on the risk of GBS, possibly due to the waning vaccine-induced immunity. The effect across different follow-up periods is shown in Supplementary Figure 2. Furthermore, the first vaccine dose was associated with a reduced risk of GBS (IRR = 0.39; 95% CI, 0.22–0.70; *P* = 0.001, Supplementary Figure 3), while the second dose was associated with an even lower risk (IRR = 0.21; 95% CI, 0.11–0.40; *P* < 0.0001, Supplementary Figure 3), both compared with the unexposed control group (Figure 3). In addition, a significant difference was observed between the first and second doses (Figure 3d). These findings suggest a potential dose-dependent association between COVID-19 vaccination and lower GBS risk during the 30-day follow-up period.

**Figure 3:**
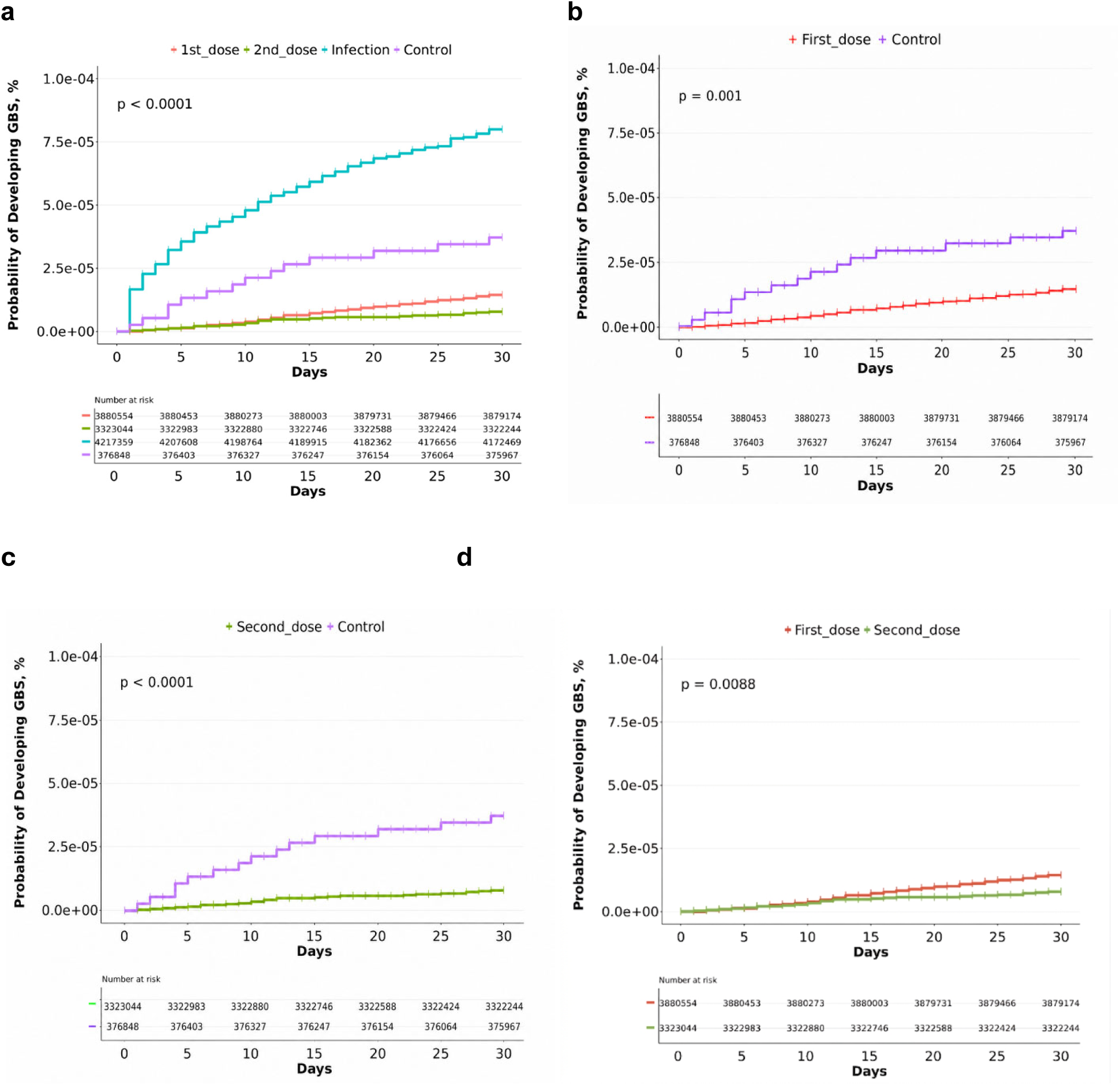
Association of COVID-19 Vaccination Doses with Risk of GBS During the 30-Day Follow-Up Period. (a) Kaplan–Meier curves representing the 30-day cumulative incidence of GBS following the first and second COVID-19 vaccine doses (b–c) Comparative analysis of the cumulative incidence of GBS within 30 days following the first (b) or second (c) dose of COVID-19 vaccination compared with the control group. (d) Cumulative curves representing GBS diagnosis within 30 days following the first and second doses of COVID-19 vaccination. The x-axis represents days from index date, and the y-axis indicates the probability of developing GBS as a percentage. Statistical significance is represented by Log-rank *P*-values. Abbreviation: GBS, Guillain-Barré Syndrome.

### Risk of GBS Associated with Different COVID-19 Vaccine Platforms

Within the N3C data Enclave, three primary COVID-19 vaccines were administered: the Johnson & Johnson/Janssen Ad26.COV2.S adenoviral vector vaccine, the Pfizer-BioNTech BNT162b2 mRNA vaccine, and the Moderna mRNA-1273 vaccine. Given the heterogeneity of COVID-19 vaccine platforms, we further aimed to evaluate which vaccine(s) contributed to the observed lower risk of GBS. Our analysis revealed a significantly decreased risk of GBS following COVID-19 mRNA vaccination compared with both unexposed control and the adenoviral vector vaccination group (Figure 4; Supplementary Figures 4 and 5). In contrast, no significant difference in GBS risk was observed between the adenoviral vector vaccine and the unexposed control group (Figure 4d; Supplementary Figure 5). These findings indicate a clear difference in GBS risk by COVID-19 vaccine platform, with a significant protective association observed exclusively for mRNA vaccines. Consistently, both mRNA vaccines were associated with a significantly reduced GBS risk compared with unexposed control: the Pfizer-BioNTech vaccine (IRR = 0.38, 95% CI, 0.20–0.71, *P* = 0.00023) and the Moderna vaccine (IRR = 0.24, 95% CI, 0.10–0.55, *P* = 0.0018) (Figure 4; Supplementary Figure 5).

**Figure 4:**
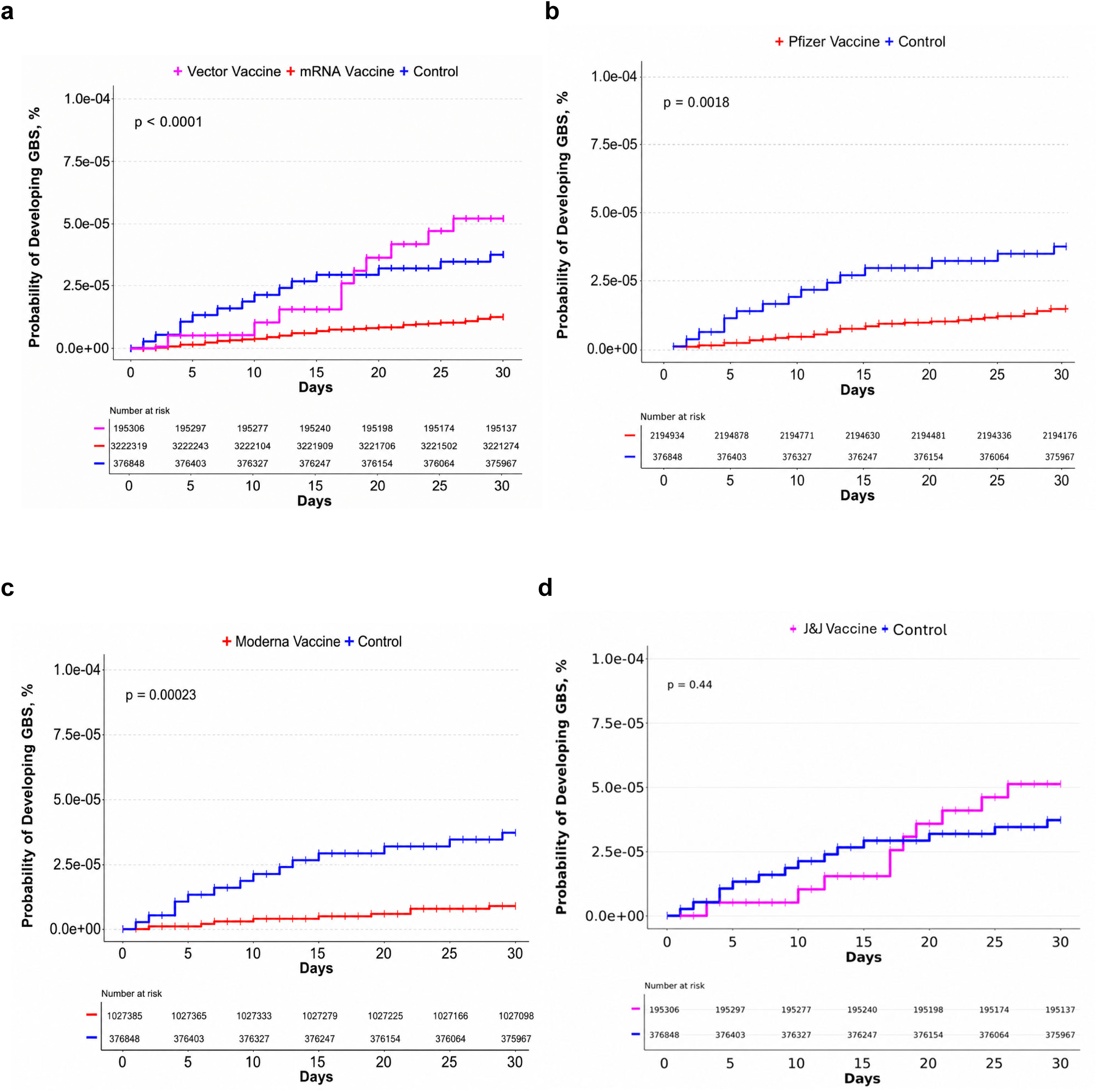
Association of GBS Risk with Different COVID-19 Vaccine Platforms and Manufacturers. (a) Kaplan–Meier curves representing the cumulative incidence of GBS within 30 days following COVID-19 mRNA or adenoviral vector vaccination. (b) Cumulative incidence of GBS following COVID-19 Pfizer mRNA vaccination compared with unexposed control. (c) Cumulative incidence of GBS following COVID-19 Moderna mRNA vaccination compared with unexposed control. (d) Cumulative incidence of GBS following adenoviral vector vaccination (J&J) compared with unexposed control. The x-axis represents days from index date, and the y-axis indicates the probability of developing GBS as a percentage. Statistical significance is represented by Log-rank *P*-values. Abbreviation: GBS, Guillain-Barré Syndrome.

**Figure 5:**
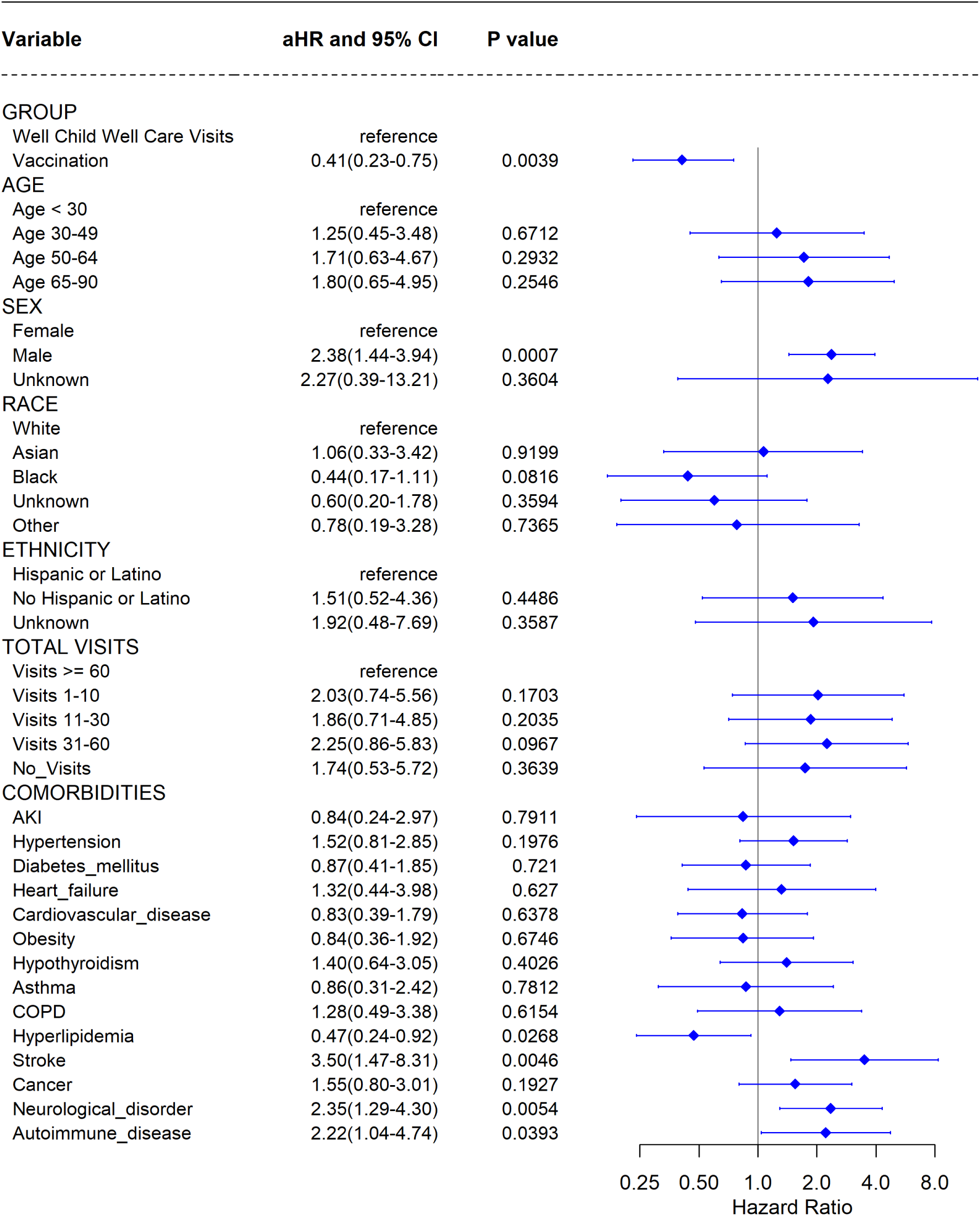
Adjusted Hazard Ratio of GBS Risk Following COVID-19 Vaccination. The multivariable Cox proportional hazards (PH) model adjusted for age, gender, race, ethnicity, total healthcare visits and comorbidities, was used to compare the GBS risk between the vaccination and control groups. Study participants were categorized into two groups based on their initial COVID-19 vaccination status during the period from December 2020 to November 2024. Each row in the table corresponds to an independent outcome, with the primary outcome evaluated over a 30-day follow-up period. Abbreviations: aHR, Adjusted hazard ratio; CI, confidence interval.

To further control for potential confounding factors from co-administration of other vaccines, such as the HPV and influenza vaccines, we conducted a sensitivity analysis by excluding individuals who received co-administration of other vaccines alongside the COVID-19 vaccine. In this analysis, the COVID-19 vaccination alone was associated with a significantly reduced risk of GBS (IRR = 0.40, 95% CI, 0.22-0.72, *P* = 0.0012, Supplementary Figures 3, 6). These findings were consistent with the primary analysis with a slightly lower IRR after excluding individuals who received co-administered other vaccines.

### Association of Prior COVID-19 Vaccination with the Risk of Infection-Related GBS

Given the observed association between COVID-19 infection and increased GBS risk, we next evaluated whether prior vaccination mitigated GBS risk among infected individuals. Prior COVID-19 vaccination within 90 days before infection was associated with a significantly lower incidence of GBS compared with COVID-19 infection without prior vaccination (IRR = 0.38, 95% CI: 0.20–0.74; *P* = 0.0031), suggesting a potential protective effect. Similar protective associations were observed among individuals vaccinated within 180 days (IRR = 0.45, 95% CI: 0.29–0.70; *P* = 0.0002) and within 365 days before COVID-19 infection (IRR = 0.43, 95% CI: 0.31–0.59; *P* < 0.0001). No significant differences in GBS risk were observed among the vaccination groups across the different time intervals examined. The effect across different prior-vaccination time periods is shown in Supplementary Figure 7. These findings suggest that COVID-19 vaccination substantially reduces the risk of infection-associated GBS, with protective effects lasting beyond the period of peak antibody-mediated immunity, consistent with previous reports.^31^

### Other Factors Associated with GBS Risk

The multivariate Cox model showed there were several comorbid conditions independently associated with increased risk of GBS, such as stroke, neurological disorders, and autoimmune diseases when we compare the COVID-19 vaccination vs. control groups (Figure 5). Moreover, males exhibited a higher risk compared to females, which is consistent with previous research.^32^ No significant associations were observed for age, racial groups and different clinical visits in the multivariable analysis. In the comparison between COVID-19 infection and control groups, individuals aged 50-90 had a significantly higher GBS risk compared to those under 30 (Supplementary Figure 8). Other variables showed similar associations as observed in the vaccination versus control groups.

## Discussion

In this large EHR-based cohort study, we found COVID-19 infection was associated with increased GBS risk, whereas COVID-19 mRNA vaccination, but not adenovector-based vaccination, was associated with a lower GBS risk compared with unexposed control. The observed temporal and dose-dependent relationship suggest a potential causal association between mRNA COVID-19 vaccination and reduced GBS risk.^33,34^

Our study is the first large-scale, systematically designed cohort study to provide robust evidence supporting a protective association between COVID-19 mRNA vaccination and the occurrence of GBS. Unlike previous studies, which primarily relied on comparisons among different COVID-19 vaccine platforms or historical control groups,^22–26^ our study incorporated concurrent control group and excluded GBS cases related to COVID-19 infection.^10–13^ This approach more effectively controlled for background GBS incidence and differences in vaccine effectiveness against COVID-19 infection, thereby reducing bias arising from differential infection risk across vaccination groups and the associated risk of infection-related GBS. As a result, it offers a more reliable assessment of the relationship between COVID-19 vaccination and GBS occurrence. On the other hand, GBS is a rare neurological disorder that occurs naturally in the population,^7^ which means passive surveillance systems can mistakenly attribute coincidental post-vaccination GBS cases to the vaccine itself. By incorporating a contemporary control group, our study was able to account for the background incidence of GBS in pandemic, allowing for a more accurate assessment of the independent effects of both COVID-19 infection and vaccination on GBS risk.

Furthermore, we found that prior COVID-19 vaccination markedly reduced the risk of GBS among individuals who later became infected with SARS-CoV-2, compared to infected individuals without prior vaccination. This protective association persisted within a year before infection, which is consistent with previous observations demonstrating that COVID-19 vaccination continues to reduce the risk of post-acute and immune-mediated sequelae following breakthrough COVID-19 infection, even as protection against infection itself wanes.^31^These results suggest that COVID-19 mRNA vaccines may offer dual protection by preventing COVID-19 infection, and consequently reducing associated GBS risk.^8^

Our analyses showed that recipients of both BNT162b2 and mRNA-1273 had a significantly lower risk of GBS compared with both adenoviral vector vaccine recipients and unexposed control groups. Consistent with our observations, emerging evidence suggests that COVID-19 mRNA vaccines may confer a protective effect against GBS, potentially through reducing the risk or severity of COVID-19 infection.^26^ A nested case-control study from Israel found that COVID-19 infection significantly increased the risk of GBS, whereas the Pfizer-BioNTech mRNA vaccine BNT162b2 appeared to be associated with a lower risk of GBS, although the association did not reach statistical significance.^26^ Similar to our study, that nested case-control study excluded COVID-19 infection as a confounder when evaluating the association between vaccination and GBS risk. In contrast, prior investigations reporting no alterations in GBS risk after COVID-19 mRNA vaccination failed to account for COVID-19 infection status as a confounder, likely due to limited clinical information availability in the early stage of pandemic.^25^ As a result, the true safety differences among COVID-19 vaccines may have been obscured by confounding from SARS-CoV-2 infection.^22,23^

While a statistically significant association between recombinant viral vector vaccine Ad26.COV2.S and incident GBS within the 30-day risk window was not found, we observed a unique dynamic trajectory of GBS occurrence during this period. Specifically, risk among Ad26.COV2.S recipients initially remained below that of the control group, crossed above it at around day 18, and remained elevated through day 30. The difference on day 30 did not reach statistical significance, likely owing to the limited sample numbers. Previous studies of Ad26.COV2.S have reported elevated GBS risk at day 21 post-vaccination, but those studies have relied on fixed observation time windows (e.g., day 21 or day 42) rather than tracking cumulative incidence over time, which may limit their ability to capture how GBS risk develops over time.^22–24^ Additionally, these studies did not exclude COVID-19 infection–related GBS cases. Given that the Janssen vaccine showed lower initial efficacy than mRNA platforms,^2–4^ recipients of Ad26.COV2.S vaccine were likely to have a higher vulnerability to breakthrough COVID-19 infections as immunity waned over time than those of COVID-19 mRNA vaccines. Thus, our study is the first to longitudinally evaluate this risk, revealing a previously unrecognized delayed crossover pattern in GBS risk following Ad26.COV2.S vaccination.

The underlying biological mechanism by which COVID-19 mRNA vaccines may protect the vaccinees against GBS is unclear.^35,36^ Our findings suggest that multiple factors may contribute to this protective association. Prior COVID-19 vaccination was associated with reduced COVID-19 infection associated GBS risk. In addition, sex and several pre-existing comorbidities, including stroke, neurological disorders and autoimmune diseases, were independently associated with GBS risk, suggesting that both demographic and clinical factors contribute to susceptibility following vaccination. Experimental studies suggest that the SARS-CoV-2 spike protein alone is unlikely to induce GBS, as repeated exposure in C57BL/6 mice did not produce detectable autoimmune responses.^37^ The absence of a consistent association between GBS and all COVID-19 vaccine platforms further weakens the hypothesis that the spike protein itself is the primary driver of disease. Instead, differences in innate and adaptive immune activation across vaccine platforms have been proposed as a more plausible explanation for heterogeneous findings. In particular, adenoviral vector vaccines may elicit strong innate and adaptive immunogenicity, which has been hypothesized to contribute to immune-mediated neurological events in susceptible individuals.^38–41^ In contrast, mRNA vaccines incorporate modified nucleosides, such as N1-methylpseudouridine, together with optimized delivery systems that enhance mRNA stability and antigen expression while attenuating innate immune sensing, thereby promoting robust adaptive immunity with reduced inflammatory activation.^42–46^ These immunological differences may partly account for the lower-than-expected incidence of GBS observed following COVID-19 mRNA-based vaccination. We did not observe an increased risk of GBS associated with adenoviral vector vaccination.

Further studies are needed to elucidate underlying mechanisms. COVID-19 vaccination may also indirectly reduce GBS incidence by decreasing SARS-CoV-2 transmission and preventing infection-associated GBS.^8^

### Strengths and Limitations

Our person-level, time-to-event design enabled continuous risk assessment, overcoming the limitations of static, aggregate reporting and yielding a more accurate and comprehensive profile of the association between COVID-19 vaccination and GBS risk. By leveraging the large, nationally representative N3C Data Enclave, which harmonizes EHR data via the OMOP Common Data Model, we minimized reporting bias and denominator uncertainty that commonly affect passive pharmacovigilance systems such as VAERS, enabling robust estimation of absolute and relative risks that such systems cannot provide.^22,47^ To better isolate vaccine-specific effects, we excluded individuals with documented COVID-19 infection from the vaccination cohorts. This exclusion is particularly important given that COVID-19 infection itself is a recognized trigger for GBS, and that adenovirus-vectored vaccines initially showed lower efficacy against infection than mRNA platforms (approximately 52.9% vs. 94–95%, respectively).^2–4^ Higher post-vaccination infection rates among adenovector recipients may therefore have confounded previous surveillance findings. Our use of a concurrent control group further strengthens this isolation of vaccine-specific risk, although differences in population structure and vaccine rollout timing may also contribute to the discrepancies observed across studies. Finally, we conducted stringent sensitivity analyses, including exclusion of recent non-COVID-19 vaccinations and adjustment for demographic and clinical covariates. Together, these analyses support the reliability and generalizability of our findings regarding the potential protective association of COVID-19 mRNA vaccination against GBS.

There were several limitations. First, the N3C Enclave data may include inaccuracies, missing values, misclassifications in COVID-19 infection or vaccination coding, or incomplete records. To mitigate potential misclassification of infection status, we included a pre-pandemic control group to help exclude undiagnosed COVID-19 and unvaccinated cases. We also adjusted for the number of hospital visits to account for variations in healthcare utilization and engagement with the N3C network. Second, the retrospective design of the study limits its ability to establish causal relationships from the observed associations. To address this issue, we performed time- and dose-related analyses, which may suggest a causal association.^33,34^ Third, the smaller number of Ad26.COV2.S limited statistical power, a challenge also reflected with wide confidence intervals in prior work.^23,24^ Finally, differences between individuals receiving the first and second vaccine doses, particularly the exclusion of those who may have developed GBS after the first dose, could introduce survivor bias.^48^

In conclusion, our study indicates that COVID-19 mRNA vaccination is associated with a lower risk of GBS, whereas adenoviral vector vaccination shows no significant change of GBS risk relative to unexposed controls. This suggests that the protective effect against GBS development may be specific to the COVID-19 mRNA vaccine platform, the mechanisms of which warrant further investigation.

## Methods

### Data Sources and Study Population

Data for this study were sourced from the National COVID Cohort Collaborative (N3C) Data Enclave, a centralized clinical repository. As of Nov. 1, 2024, N3C contained longitudinal records for over 22.8 million patients across 98 sites in the United States. ^49,50^ Data partner sites contribute demographic, visit, vital status, medication, laboratory, diagnoses, pharmacy records, both inpatient administrations and outpatient prescriptions to a central data repository. The data is harmonized from diverse sources, such as ACT, PCORnet, and TriNetX, on a regular basis according to the Observational Medical Outcomes Partnership (OMOP) common data model (CDM), which enable standardization of the N3C vocabularies for consistent data integration, analysis, and sharing in advanced clinical research.^51–53^ The N3C is supported by the National Center for Advancing Translational Science (NCATS) of the National Institutes of Health (NIH), with data and supporting analytics are hosted on the N3C Enclave data platform. Data transferred to the NCATS from N3C is governed by a Johns Hopkins University Reliance Protocol (IRB00249128) or through individual data partner agreements with the NIH. Data usage for this study was authorized by N3C (DUR-36ED2AE) and reviewed and approved by the Medical School Institutional Review Board (IRB) at the University of Michigan (HUM00243962). All data usage was in compliance with N3C’s data use policies and obligations. For detailed data ingestion, quality control and more, please refer to Supplementary Methods.

### Study Design and Cohort Building

This retrospective cohort study^54^ utilized de-identified N3C data from December 11, 2020 (the introduction of COVID-19 vaccines) through November 1, 2024. The patient recruitment process and study design were depicted in Figure 1 and 6.

**Figure 6:**
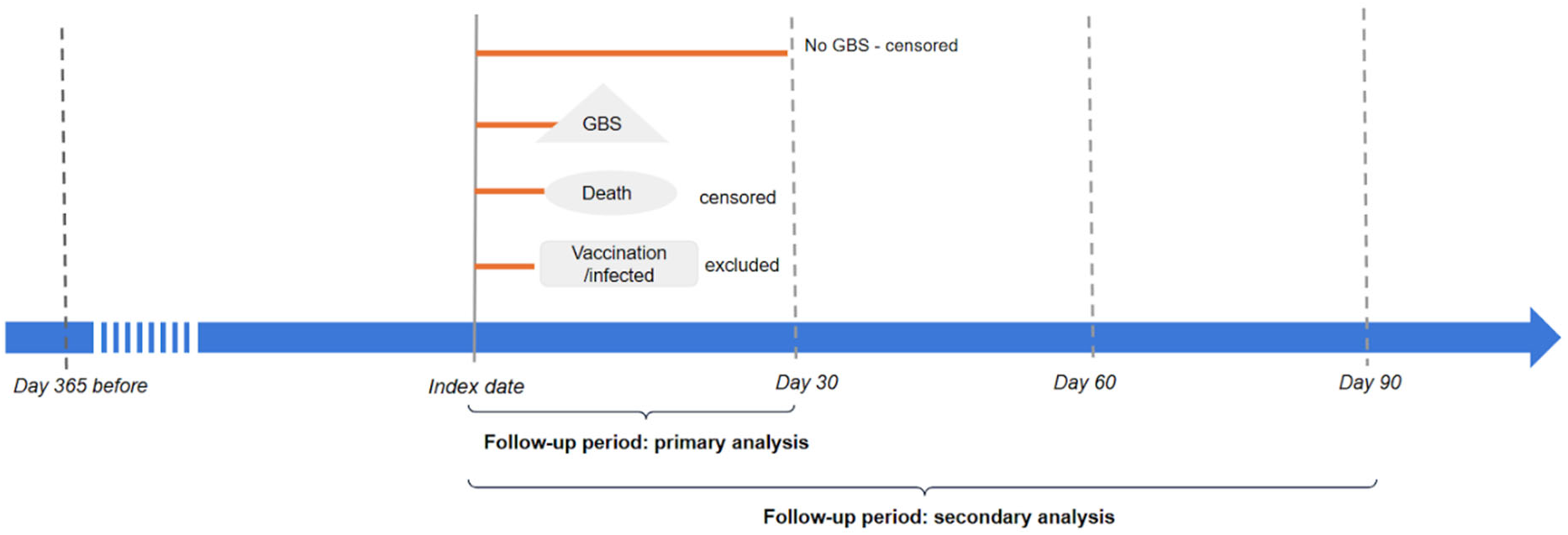
Study Design for Evaluating the Association Between GBS diagnosis and COVID-19 Vaccination or Infection. The diagram outlines the study period, starting from one year before the index date and extending to 90 days post-exposure. The initial occurrence of vaccination, infection or well-care visit was defined as the index event. Participants were followed for 30 days in the primary analysis, with follow-up extended to 60 or 90 days in the secondary analyses. Individuals were censored at the end of the respective follow-up period if no GBS events occurred or upon death within 30, 60, or 90 days. A one-year look-back period before the index date was used to assess demographics and comorbidities, ensuring the inclusion of only new-onset events. Participants with a diagnosis of GBS within 365 days prior to the index date were excluded from the analysis. Abbreviation: GBS, Guillain-Barré Syndrome.

Three mutually exclusive groups were established: (1) COVID-19 vaccination, (2) COVID-19 infection, and (3) unexposed well-care visit control (a commonly method for defining of control group in COVID-19 studies in N3C data Enclave).^55^ Participants were assigned to the vaccination group if their first recorded COVID-19 vaccination occurred before any documented COVID-19 infection and no COVID-19 infection occurred during the follow-up period; the index date was defined as the date of first vaccination. Conversely, participants were assigned to the infection group if their first documented COVID-19 infection occurred before any COVID-19 vaccination and no vaccination was received during the follow-up period; the index date was defined as the date of first infection. Participants were assigned to the unexposed control group if they had no record of COVID-19 vaccination or infection before the index visit or during the follow-up period; the index date was defined as the date of their first well-care visit.^55^ The index date for the vaccination group was set as the date of the first dose vaccination to evaluate early immune responses potentially associated with GBS, while reducing selection bias by including individuals who might not have completed the vaccination series due to adverse events. For the control group, the index date was determined based on well-care visit date, which was unrelated to COVID-19 vaccination or infection, as previously described.^55^ Cohort-specific inclusion and exclusion criteria were applied (Figure 1). Patients with a diagnosis of GBS within 365 days before the index date were excluded to reduce misclassification of prevalent or recent prior GBS as incident events during follow-up. For the vaccination group, a secondary analysis was conducted using the date of the second vaccine dose as the index date. Participants were assigned to the second dose vaccination group if their second recorded COVID-19 vaccination occurred before any documented COVID-19 infection and there was no infection during the follow-up; the index date was defined as the date of the second vaccination. The patient recruitment process was depicted in Figure 1.

### Exposures

The exposures of interest were COVID-19 infection and COVID-19 vaccination. Individuals with neither documented COVID-19 infection nor COVID-19 vaccination during the study period were included as an unexposed control. COVID-19 infection was identified using diagnosis codes and laboratory-confirmed COVID-19 test results, including viral culture and nucleic acid amplification assays. COVID-19 vaccination data were collected between December 11, 2020, the date of the U.S. COVID-19 vaccine rollout, and November 1, 2024, the end of the study period. The vaccines evaluated included the Johnson & Johnson adenoviral vector vaccine, the Pfizer-BioNTech and Moderna mRNA vaccines. Vaccination records were identified with related concept ids of OMOP CDM from the N3C data Enclave (Supplementary Table 1).

### Outcome

Incidence of GBS during the follow-up period after the index date was identified using standardized OMOP concept IDs (Supplementary Table 1). A structured query of the N3C database was conducted to identify GBS and related variants based on predefined MedDRA preferred terms (PTs), including acute motor axonal neuropathy, acute motor-sensory axonal neuropathy, autoimmune neuropathy, demyelinating polyneuropathy, demyelination, subacute inflammatory demyelinating polyneuropathy, immune-mediated neuropathy, Guillain-Barré syndrome, and Miller-Fisher syndrome (Supplementary Table 1).

### Statistical Analysis

We assessed each patient’s comorbidities within one year prior to the index date and summarized cohort characteristics according to demographic variables and pre-existing comorbid conditions. Differences in demographic characteristics and GBS incidence were evaluated using chi-square tests for categorical variables and two-sample t-tests for continuous variables. Absolute risk rate and relative incidence risk ratio of GBS along with their 95% confidence intervals were calculated. Kaplan-Meier estimates were used to assess the probability of developing GBS within the 30-day follow-up period and to visualize the differences across exposure groups.^56^ Differences in these curves were tested using the Log-rank test. Study subjects were followed from the index date until the earliest occurrence of GBS, death from any cause, or end of the 30 days follow-up window (Figure 6). We also conducted secondary analyses extending the study period to 60 days and 90 days. A multivariable Cox proportional hazards regression model, commonly used to assess GBS risk,^57–60^ was applied to estimate adjusted hazard ratios for GBS, adjusting for baseline demographic characteristics and comorbidities previously reported to be associated with COVID-19 related GBS risk.^61^

Due to concerns that some individuals in the pandemic-era control group may have experienced COVID-19 infection that was not captured in the EHR (e.g., asymptomatic infection, home testing), potentially biasing the estimated associations, we additionally included a pre-pandemic control group in which COVID-19 exposure was not possible to assess the robustness of the observed associations. A pre-pandemic control group (consisting of patients with well-care visits from January 30, 2018, to January 30, 2020) was included for a sensitivity analysis.

## Data Availability

All data of this study is available in the N3C Data Enclave for researchers with an approved protocol and data use request from an institutional review board. Data access is governed by the National Institutes of Health. More information on the enclave and instructions for data access can be found at https://covid.cd2h.org/for-researchers.

## Code Availability

All statistical analysis code has been deposited in a publicly accessible OSF repository at: https://github.com/lijinju/COVID-19-Vaccine-and-Guillain-Barre-Syndrome

## Supplementary Material 1

Supplementary Methods: Detailed Characterization, Quality Assurance, and Harmonization of the N3C Data Enclave

Supplementary Information of N3C Contributors and Partners: Core contributors to N3C and Data Partners with Released Data

## Supplementary Material 2

Supplementary Table 1 a, b, c, d, e: Concept IDs Used to Define COVID-19 Vaccination, COVID-19 Infection with Diagnostic, COVID-19 Infection with Lab Test, Well-Care Visit, and GBS Incidence in N3C Enclave

Supplementary Figure 1: Forest Plot of Incidence Risk Ratios for GBS After COVID-19 Vaccination or Infection During 30-Day Follow-up

Supplementary Figure 2: Association Between COVID-19 Vaccination and GBS Incidence Across Post-Vaccination Follow-Up Periods Using a Multivariable-Adjusted Cox Model

Supplementary Figure 3: Risk of GBS Within 30 Days After COVID-19 Vaccination Stratified by Doses or Co-administered Vaccines

Supplementary Figure 4: Comparison of GBS Risk Following COVID-19 mRNA and Adenoviral Vector Vaccination During 30-Day Follow-up Period

Supplementary Figure 5: Association Between COVID-19 Vaccine Manufacturers and Risk of GBS Within 30 Days of Vaccination

Supplementary Figure 6: Association of GBS Risk Within 30 Days Following COVID-19 Vaccination After Excluding Other Vaccine Coadministration

Supplementary Figure 7: Risk of GBS Following COVID-19 Infection Stratified by Pre-Infection Vaccination-to-Infection Time Interval

Supplementary Figure 8: Association Between COVID-19 Infection and GBS Risk Evaluated Using a Multivariable-Adjusted Cox Proportional Hazards Model

## Acknowledgements

This study was supported by two grants, U24AI171008 (Y.He) and National Institute of Allergy and Infectious Diseases of the National Institutes of Health under award number R01AI158543 (L.Z. and Y.He). The funders played no role in the study design, data collection, analysis, and interpretation of data, or the writing of this manuscript.

## N3C Attribution

The analyses described in this report were conducted with data or tools accessed through the NCATS N3C Data Enclave https://covid.cd2h.org and N3C Attribution & Publication Policy v 1.2-2020-08-25b supported by NCATS Contract No. 75N95023D00001, Axle Informatics Subcontract: NCATS-P00438-B. This research was possible because of the patients whose information is included within the data and the organizations (https://ncats.nih.gov/n3c/resources/data-contribution/data-transfer-agreement-signatories) and scientists who have contributed to the on-going development of this community resource [https://doi.org/10.1093/jamia/ocaa196].

For a complete list of additional N3C Core Contributors and Data Partners with released data, please refer to the Supplementary Materials.

## Author Contributions

JL and YHe initiated the project and provided the original project design. JL was responsible for data analysis and writing the first version of the manuscript. JL and YP were responsible for cohort data generation. JL, YHan, CZ, LZ, YH developed research questions and ways to address the questions. LZ and CZ served as statistics experts. YHe served as the vaccine adverse event expert. All authors participated in result interpretation, discussion, and paper editing. The corresponding author is YHe.

## Consortia Authorship

Christopher G. Chute^10^

^10^Schools of Medicine, Public Health, and Nursing, Johns Hopkins University, Baltimore, MD, USA

## Disclaimer

The N3C Publication committee confirmed that this manuscript is in accordance with N3C data use and attribution policies; however, this content is solely the responsibility of the authors and does not necessarily represent the official views of the National Institutes of Health or the N3C program.

