## Supplementary Material 1 for "COVID-19 mRNA Vaccination Reduces Guillain-Barré Syndrome Risk: Evidence from a Large Longitudinal Cohort Study"

Supplementary Methods: Detailed Characterization, Quality Assurance, and Harmonization of the N3C Data Enclave

Supplementary Information of N3C Contributors and Partners: Core contributors to N3C and Data Partners with Released Data

**Supplementary Methods: Characterization, Quality Assurance, and Harmonization of the N3C Data Enclave**

**Overview of the N3C Data Enclave**

This study utilized the National COVID Cohort Collaborative (N3C) Data Enclave, a sophisticated, secure platform hosted by the NIH’s National Center for Advancing Translational Sciences (NCATS). Beyond being a simple repository, the N3C functions as a dynamic ecosystem that aggregates and harmonizes Electronic Health Records (EHR) from 98 distinct clinical organizations across the U.S at the time of the study conducting.^1^ To solve the inherent heterogeneity of electronic health record (EHR) data, N3C harmonized disparate source datasets from contributing sites into the Observational Medical Outcomes Partnership (OMOP) Common Data Model (CDM).^2,3^ This standardization enables comprehensive characterization of the patient journey, including outpatient encounters, emergency department visits, hospitalizations, and intensive care stays. The repository is vast, containing longitudinal data for over 22 million patients, which includes detailed clinical information, including demographics, healthcare encounters, diagnoses, procedures, medications, vital signs, laboratory results, vaccinations, and other relevant clinical observations.^1-4^

The N3C technical infrastructure is built on a FedRAMP Moderate cloud environment, ensuring high-level security while providing researchers access to the Foundry data science platform. This platform leverages Apache Spark to handle distributed computing, enabling the processing of "big data" that would be impossible on standard hardware. Because the enclave supports various programming languages including Python, R, SQL, and Java, it fosters a community-driven environment where code and workflows can be shared and reproduced. This transparency is critical for COVID-19 research, as it allows the scientific community to validate findings and build upon existing models in real-time. ^4^

**Data Ingestion and Harmonization**

Data collection for the N3C is conducted under approval from the NIH Institutional Review Board (IRB00249128), with the Johns Hopkins University School of Medicine serving as the central IRB. Institutional review boards at participating data partner sites either approved the protocol locally or ceded oversight to the central IRB. All contributing sites have executed Data Transfer Agreements with NCATS and maintain appropriate IRB approval to support data contribution to N3C. The data originates from routine clinical care and is contributed to the N3C under a HIPAA waiver. Partner institutions submit data using one of four common data models (CDMs): OMOP, PCORnet, TriNetX, or ACT. Submitted datasets undergo model-specific quality assessments before being harmonized into the OMOP version 5.3.1 CDM, followed by additional cross-site data quality checks prior to release. Harmonization includes mapping source vocabularies (e.g., ICD-10, SNOMED) to standardized concepts within the OMOP framework. Data are ingested daily and released to the Enclave weekly. Patient records include a wide range of clinical information structured according to the OMOP CDM. ^4^ Additional information on the OMOP CDM is available through the Observational Health Data Sciences and Informatics (OHDSI) program documentation (<https://www.ohdsi.org/data-standardization/the-common-data-model/>) or OMOP GitHub repository (<https://ohdsi.github.io/CommonDataModel/>).

**Data Quality Control and Characterization**

To ensure high data accuracy and reliability, N3C implements a rigorous, multi-tiered data quality control process. Each site's initial submission is first loaded into the N3C Data Quality Portal (DQP), where automated data quality (DQ) checks are performed before manual review by the Data Ingestion & Harmonization (DI&H) team. At the contributing sites, extraction scripts check for duplicate primary keys; any duplicates cause the extraction to fail until resolved. Upon successful extraction, a data manifest containing site and payload metadata is generated, and the payload is securely transferred via sFTP to N3C for ingestion.^5^

During ingestion, the payload undergoes initial validation against formatting requirements and the manifest. It is then loaded into a database mirroring the source's native CDM to confirm conformance. A series of automated DQ checks, including COVID-19-specific code validations, are executed, with minimal corrections applied as needed (documented in the payload). The data is subsequently transformed to OMOP 5.3.1 using validated mapping rules from the native CDM. Post-transformation, a subset of the Observational Health Data Sciences and Informatics (OHDSI) Data Quality Dashboard (DQD) tests, running thousands of automated checks per site ingest, are applied to assess plausibility, conformance, and completeness, flagging issues such as outliers or temporal inconsistencies. Results are documented and added to the payload. The harmonized payload is then merged into a centralized database with all prior site data, undergoing final conformance checks before export to the analytics pipeline. To minimize data loss and preserve contextual details for downstream meta-analyses, raw source data are retained throughout mapping and transformation. This multi-layered approach, combining automated tools like the DQD, manual oversight, and standardized transformation maintains high-fidelity data suitable for robust, large-scale COVID-19 research.^4^

**N3C Cohort Identification**

Cohort identification within the N3C repository relies on OMOP Concept Identifiers (Concept IDs) mapped from patients’ health records, including diagnoses, medications, procedures, laboratory results, and vaccination administrations. These Concept IDs harmonize a wide variety of source vocabularies submitted by participating institutions, such as ICD-10, CPT, RxNorm, and SNOMED CT, into standardized, unique OMOP identifiers, enabling consistent and reliable capture of clinical data across sites.^6^ Defining a cohort begins with the development of a computable phenotype, which leverages these OMOP Concepts and organizes them into Concept Sets (collections of one or more related Concepts representing a clinical criterion). Using the ATLAS tool, source codes are translated into OMOP Concepts, grouped into Concept Sets, and then applied to identify patients who meet the phenotype criteria. This process ensures robust, reproducible, and inclusive cohort creation, supporting high-quality analyses across the harmonized OMOP dataset in the N3C repository.

Core contributors to N3C:

Adam B. Wilcox, Adam M. Lee, Alexis Graves, Alfred (Jerrod) Anzalone, Amin Manna, Amit Saha, Amy Olex, Andrea Zhou, Andrew E. Williams, Andrew M. Southerland, Andrew T. Girvin, Anita Walden, Anjali Sharathkumar, Benjamin Amor, Benjamin Bates, Brian Hendricks, Brijesh Patel, G. Caleb Alexander, Carolyn T. Bramante, Cavin Ward-Caviness, Charisse Madlock-Brown, Christine Suver, Christopher G. Chute, Christopher Dillon, Chunlei Wu, Clare Schmitt, Cliff Takemoto, Dan Housman, Davera Gabriel, David A. Eichmann, Diego Mazzotti, Donald E. Brown, Eilis Boudreau, Elaine L. Hill, Emily Carlson Marti, Emily R. Pfaff, Evan French, Farrukh M Koraishy, Federico Mariona, Fred Prior, George Sokos, Greg Martin, Harold P. Lehmann, Heidi Spratt, Hemalkumar B. Mehta, J.W. Awori Hayanga, Jami Pincavitch, Jaylyn Clark, Jeremy Richard Harper, Jessica Yasmine Islam, Jin Ge, Joel Gagnier, Johanna J. Loomba, John B. Buse, Jomol Mathew, Joni L. Rutter, Julie A. McMurry, Justin Guinney, Justin Starren, Karen Crowley, Katie Rebecca Bradwell, Kellie M. Walters, Ken Wilkins, Kenneth R. Gersing, Kenrick Cato, Kimberly Murray, Kristin Kostka, Lavance Northington, Lee Pyles, Lesley Cottrell, Lili M. Portilla, Mariam Deacy, Mark M. Bissell, Marshall Clark, Mary Emmett, Matvey B. Palchuk, Melissa A. Haendel, Meredith Adams, Meredith Temple-O'Connor, Michael G. Kurilla, Michele Morris, Nasia Safdar, Nicole Garbarini, Noha Sharafeldin, Ofer Sadan, Patricia A. Francis, Penny Wung Burgoon, Philip R.O. Payne, Randeep Jawa, Rebecca Erwin-Cohen, Rena C. Patel, Richard A. Moffitt, Richard L. Zhu, Rishikesan Kamaleswaran, Robert Hurley, Robert T. Miller, Saiju Pyarajan, Sam G. Michael, Samuel Bozzette, Sandeep K. Mallipattu, Satyanarayana Vedula, Scott Chapman, Shawn T. O'Neil, Soko Setoguchi, Stephanie S. Hong, Steven G. Johnson, Tellen D. Bennett, Tiffany J. Callahan, Umit Topaloglu, Valery Gordon, Vignesh Subbian, Warren A. Kibbe, Wenndy Hernandez, Will Beasley, Will Cooper, William Hillegass, Xiaohan Tanner Zhang. Details of contributions available at covid.cd2h.org/core-contributors

Data Partners with Released Data

The following institutions whose data is released or pending:

Available: Advocate Health Care Network — UL1TR002389: The Institute for Translational Medicine (ITM) • Aurora Health Care Inc — UL1TR002373: Wisconsin Network For Health Research • Boston University Medical Campus — UL1TR001430: Boston University Clinical and Translational Science Institute • Brown University — U54GM115677: Advance Clinical Translational Research (Advance-CTR) • Carilion Clinic — UL1TR003015: iTHRIV Integrated Translational health Research Institute of Virginia • Case Western Reserve University — UL1TR002548: The Clinical & Translational Science Collaborative of Cleveland (CTSC) • Charleston Area Medical Center — U54GM104942: West Virginia Clinical and Translational Science Institute (WVCTSI) • Children’s Hospital Colorado — UL1TR002535: Colorado Clinical and Translational Sciences Institute • Columbia University Irving Medical Center — UL1TR001873: Irving Institute for Clinical and Translational Research • Dartmouth College — None (Voluntary) Duke University — UL1TR002553: Duke Clinical and Translational Science Institute • George Washington Children’s Research Institute — UL1TR001876: Clinical and Translational Science Institute at Children’s National (CTSA-CN) • George Washington University — UL1TR001876: Clinical and Translational Science Institute at Children’s National (CTSA-CN) • Harvard Medical School — UL1TR002541: Harvard Catalyst • Indiana University School of Medicine — UL1TR002529: Indiana Clinical and Translational Science Institute • Johns Hopkins University — UL1TR003098: Johns Hopkins Institute for Clinical and Translational Research • Louisiana Public Health Institute — None (Voluntary) • Loyola Medicine — Loyola University Medical Center • Loyola University Medical Center — UL1TR002389: The Institute for Translational Medicine (ITM) • Maine Medical Center — U54GM115516: Northern New England Clinical & Translational Research (NNE-CTR) Network • Mary Hitchcock Memorial Hospital & Dartmouth Hitchcock Clinic — None (Voluntary) • Massachusetts General Brigham — UL1TR002541: Harvard Catalyst • Mayo Clinic Rochester — UL1TR002377: Mayo Clinic Center for Clinical and Translational Science (CCaTS) • Medical University of South Carolina — UL1TR001450: South Carolina Clinical & Translational Research Institute (SCTR) • MITRE Corporation — None (Voluntary) • Montefiore Medical Center — UL1TR002556: Institute for Clinical and Translational Research at Einstein and Montefiore • Nemours — U54GM104941: Delaware CTR ACCEL Program • NorthShore University HealthSystem — UL1TR002389: The Institute for Translational Medicine (ITM) • Northwestern University at Chicago — UL1TR001422: Northwestern University Clinical and Translational Science Institute (NUCATS) • OCHIN — INV-018455: Bill and Melinda Gates Foundation grant to Sage Bionetworks • Oregon Health & Science University — UL1TR002369: Oregon Clinical and Translational Research Institute • Penn State Health Milton S. Hershey Medical Center — UL1TR002014: Penn State Clinical and Translational Science Institute • Rush University Medical Center — UL1TR002389: The Institute for Translational Medicine (ITM) • Rutgers, The State University of New Jersey — UL1TR003017: New Jersey Alliance for Clinical and Translational Science • Stony Brook University — U24TR002306 • The Alliance at the University of Puerto Rico, Medical Sciences Campus — U54GM133807: Hispanic Alliance for Clinical and Translational Research (The Alliance) • The Ohio State University — UL1TR002733: Center for Clinical and Translational Science • The State University of New York at Buffalo — UL1TR001412: Clinical and Translational Science Institute • The University of Chicago — UL1TR002389: The Institute for Translational Medicine (ITM) • The University of Iowa — UL1TR002537: Institute for Clinical and Translational Science • The University of Miami Leonard M. Miller School of Medicine — UL1TR002736: University of Miami Clinical and Translational Science Institute • The University of Michigan at Ann Arbor — UL1TR002240: Michigan Institute for Clinical and Health Research • The University of Texas Health Science Center at Houston — UL1TR003167: Center for Clinical and Translational Sciences (CCTS) • The University of Texas Medical Branch at Galveston — UL1TR001439: The Institute for Translational Sciences • The University of Utah — UL1TR002538: Uhealth Center for Clinical and Translational Science • Tufts Medical Center — UL1TR002544: Tufts Clinical and Translational Science Institute • Tulane University — UL1TR003096: Center for Clinical and Translational Science • The Queens Medical Center — None (Voluntary) • University Medical Center New Orleans — U54GM104940: Louisiana Clinical and Translational Science (LA CaTS) Center • University of Alabama at Birmingham — UL1TR003096: Center for Clinical and Translational Science • University of Arkansas for Medical Sciences — UL1TR003107: UAMS Translational Research Institute • University of Cincinnati — UL1TR001425: Center for Clinical and Translational Science and Training • University of Colorado Denver, Anschutz Medical Campus — UL1TR002535: Colorado Clinical and Translational Sciences Institute • University of Illinois at Chicago — UL1TR002003: UIC Center for Clinical and Translational Science • University of Kansas Medical Center — UL1TR002366: Frontiers: University of Kansas Clinical and Translational Science Institute • University of Kentucky — UL1TR001998: UK Center for Clinical and Translational Science • University of Massachusetts Medical School Worcester — UL1TR001453: The UMass Center for Clinical and Translational Science (UMCCTS) • University Medical Center of Southern Nevada — None (voluntary) • University of Minnesota — UL1TR002494: Clinical and Translational Science Institute • University of Mississippi Medical Center — U54GM115428: Mississippi Center for Clinical and Translational Research (CCTR) • University of Nebraska Medical Center — U54GM115458: Great Plains IDeA-Clinical & Translational Research • University of North Carolina at Chapel Hill — UL1TR002489: North Carolina Translational and Clinical Science Institute • University of Oklahoma Health Sciences Center — U54GM104938: Oklahoma Clinical and Translational Science Institute (OCTSI) • University of Pittsburgh — UL1TR001857: The Clinical and Translational Science Institute (CTSI) • University of Pennsylvania — UL1TR001878: Institute for Translational Medicine and Therapeutics • University of Rochester — UL1TR002001: UR Clinical & Translational Science Institute • University of Southern California — UL1TR001855: The Southern California Clinical and Translational Science Institute (SC CTSI) • University of Vermont — U54GM115516: Northern New England Clinical & Translational Research (NNE-CTR) Network • University of Virginia — UL1TR003015: iTHRIV Integrated Translational health Research Institute of Virginia • University of Washington — UL1TR002319: Institute of Translational Health Sciences • University of Wisconsin-Madison — UL1TR002373: UW Institute for Clinical and Translational Research • Vanderbilt University Medical Center — UL1TR002243: Vanderbilt Institute for Clinical and Translational Research • Virginia Commonwealth University — UL1TR002649: C. Kenneth and Dianne Wright Center for Clinical and Translational Research • Wake Forest University Health Sciences — UL1TR001420: Wake Forest Clinical and Translational Science Institute • Washington University in St. Louis — UL1TR002345: Institute of Clinical and Translational Sciences • Weill Medical College of Cornell University — UL1TR002384: Weill Cornell Medicine Clinical and Translational Science Center • West Virginia University — U54GM104942: West Virginia Clinical and Translational Science Institute (WVCTSI)  Submitted: Icahn School of Medicine at Mount Sinai — UL1TR001433: ConduITS Institute for Translational Sciences • The University of Texas Health Science Center at Tyler — UL1TR003167: Center for Clinical and Translational Sciences (CCTS) • University of California, Davis — UL1TR001860: UCDavis Health Clinical and Translational Science Center • University of California, Irvine — UL1TR001414: The UC Irvine Institute for Clinical and Translational Science (ICTS) • University of California, Los Angeles — UL1TR001881: UCLA Clinical Translational Science Institute • University of California, San Diego — UL1TR001442: Altman Clinical and Translational Research Institute • University of California, San Francisco — UL1TR001872: UCSF Clinical and Translational Science Institute  NYU Langone Health Clinical Science Core, Data Resource Core, and PASC Biorepository Core — OTA-21-015A: Post-Acute Sequelae of SARS-CoV-2 Infection Initiative (RECOVER)  Pending: Arkansas Children’s Hospital — UL1TR003107: UAMS Translational Research Institute • Baylor College of Medicine — None (Voluntary) • Children’s Hospital of Philadelphia — UL1TR001878: Institute for Translational Medicine and Therapeutics • Cincinnati Children’s Hospital Medical Center — UL1TR001425: Center for Clinical and Translational Science and Training • Emory University — UL1TR002378: Georgia Clinical and Translational Science Alliance • HonorHealth — None (Voluntary) • Loyola University Chicago — UL1TR002389: The Institute for Translational Medicine (ITM) • Medical College of Wisconsin — UL1TR001436: Clinical and Translational Science Institute of Southeast Wisconsin • MedStar Health Research Institute — None (Voluntary) • Georgetown University — UL1TR001409: The Georgetown-Howard Universities Center for Clinical and Translational Science (GHUCCTS) • MetroHealth — None (Voluntary) • Montana State University — U54GM115371: American Indian/Alaska Native CTR • NYU Langone Medical Center — UL1TR001445: Langone Health’s Clinical and Translational Science Institute • Ochsner Medical Center — U54GM104940: Louisiana Clinical and Translational Science (LA CaTS) Center • Regenstrief Institute — UL1TR002529: Indiana Clinical and Translational Science Institute • Sanford Research — None (Voluntary) • Stanford University — UL1TR003142: Spectrum: The Stanford Center for Clinical and Translational Research and Education • The Rockefeller University — UL1TR001866: Center for Clinical and Translational Science • The Scripps Research Institute — UL1TR002550: Scripps Research Translational Institute • University of Florida — UL1TR001427: UF Clinical and Translational Science Institute • University of New Mexico Health Sciences Center — UL1TR001449: University of New Mexico Clinical and Translational Science Center • University of Texas Health Science Center at San Antonio — UL1TR002645: Institute for Integration of Medicine and Science • Yale New Haven Hospital — UL1TR001863: Yale Center for Clinical Investigation
