## Supplementary Material 2 for "COVID-19 mRNA Vaccination Reduces Guillain-Barré Syndrome Risk: Evidence from a Large Longitudinal Cohort Study"

Supplementary Table 1 a, b, c, d, e: Concept IDs Used to Define COVID-19 Vaccination, COVID-19 Infection with Diagnostic, COVID-19 Infection with Lab Test, Well-Care Visit, and GBS Incidence in N3C Enclave

Supplementary Figure 1: Forest Plot of Incidence Risk Ratios for GBS After COVID-19 Vaccination or Infection During 30-Day Follow-up

Supplementary Figure 2:  Association Between COVID-19 Vaccination and GBS Incidence Across Post-Vaccination Follow-Up Periods Using a Multivariable-Adjusted Cox Model

Supplementary Figure 3: Risk of GBS Within 30 Days After COVID-19 Vaccination Stratified by Doses or Co-administered Vaccines

Supplementary Figure 4: Comparison of GBS Risk Following COVID-19 mRNA and Adenoviral Vector Vaccination During 30-Day Follow-up Period

Supplementary Figure 5: Association Between COVID-19 Vaccine Manufacturers and Risk of GBS Within 30 Days of Vaccination

Supplementary Figure 6: Association of GBS Risk Within 30 Days Following COVID-19 Vaccination After Excluding Other Vaccine Coadministration

Supplementary Figure 7: Risk of GBS Following COVID-19 Infection Stratified by Pre-Infection Vaccination-to-Infection Time Interval

Supplementary Figure 8: Association Between COVID-19 Infection and GBS Risk Evaluated Using a Multivariable-Adjusted Cox Proportional Hazards Model

**Supplementary Table 1 a, b, c, d, e: Concept IDs Used to Define COVID-19 Vaccination, COVID-19 Infection with Diagnostic, COVID-19 Infection with Lab Test, Well-Care Visit, and GBS Incidence in N3C Enclave**

| **Supplementary Table 1. a: COVID-19 Vaccines Concept IDs** | |
| --- | --- |
| **Concept IDs** | **Concept name** |
| 42649039 | MODERNA COVID-19 VACCINE, BIVALENT - moderna covid-19 vaccine, bivalent injection, suspension |
| 37003432 | SARS-CoV-2 (COVID-19) vaccine, mRNA spike protein |
| 37003434 | SARS-CoV-2 (COVID-19) vaccine, mRNA spike protein Injectable Product |
| 37003435 | SARS-CoV-2 (COVID-19) vaccine, mRNA spike protein Injectable Suspension |
| 1759205 | SARS-CoV-2 (COVID-19) vaccine, mRNA spike protein Injectable Suspension [Comirnaty] |
| 779678 | SARS-CoV-2 (COVID-19) vaccine, mRNA spike protein Injectable Suspension [Spikevax] |
| 1525539 | SARS-CoV-2 (COVID-19) vaccine, mRNA spike protein Injection |
| 722118 | SARS-COV-2 (COVID-19) vaccine, mRNA, spike protein, LNP, bivalent, preservative free, 10 mcg/0.2 mL dose |
| 722117 | SARS-COV-2 (COVID-19) vaccine, mRNA, spike protein, LNP, bivalent, preservative free, 10 mcg/0.2 mL dose, tris-sucrose formulation |
| 722119 | SARS-COV-2 (COVID-19) vaccine, mRNA, spike protein, LNP, bivalent, preservative free, 3 mcg/0.2 mL dose, tris-sucrose formulation |
| 722120 | SARS-COV-2 (COVID-19) vaccine, mRNA, spike protein, LNP, bivalent, preservative free, 30 mcg/0.3 mL dose, tris-sucrose formulation |
| 778267 | SARS-COV-2 (COVID-19) vaccine, mRNA, spike protein, LNP, bivalent, preservative free, 50 mcg/0.5 mL or 25 mcg/0.25 mL dose |
| 702678 | SARS-COV-2 (COVID-19) vaccine, mRNA, spike protein, LNP, preservative free, 10 mcg/0.2mL dose, tris-sucrose formulation |
| 724906 | SARS-COV-2 (COVID-19) vaccine, mRNA, spike protein, LNP, preservative free, 100 mcg/0.5mL dose or 50 mcg/0.25mL dose |
| 702676 | SARS-COV-2 (COVID-19) vaccine, mRNA, spike protein, LNP, preservative free, 3 mcg/0.2mL dose, tris-sucrose formulation |
| 724907 | SARS-COV-2 (COVID-19) vaccine, mRNA, spike protein, LNP, preservative free, 30 mcg/0.3mL dose |
| 702677 | SARS-COV-2 (COVID-19) vaccine, mRNA, spike protein, LNP, preservative free, 30 mcg/0.3mL dose, tris-sucrose formulation |
| 905420 | SARS-COV-2 (COVID-19) vaccine, mRNA, spike protein, LNP, preservative free, 50 mcg/0.5 mL dose |
| 778266 | SARS-COV-2 (COVID-19) vaccine, mRNA, spike protein, LNP, preservative free, pediatric 25 mcg/0.25 mL dose |
| 778265 | SARS-COV-2 (COVID-19) vaccine, mRNA, spike protein, LNP, preservative free, pediatric 50 mcg/0.5 mL dose |
| 37003516 | SARS-CoV-2 (COVID-19) vaccine, mRNA-1273 |
| 742036 | SARS-CoV-2 (COVID-19) vaccine, mRNA-1273 0.025 MG/ML |
| 42647483 | SARS-CoV-2 (COVID-19) vaccine, mRNA-1273 0.025 MG/ML / SARS-CoV-2 (COVID-19) vaccine, mRNA-1273 OMICRON (BA.4/BA.5) 0.025 MG/ML Injectable Suspension |
| 1525540 | SARS-CoV-2 (COVID-19) vaccine, mRNA-1273 0.05 MG/ML |
| 36868353 | SARS-CoV-2 (COVID-19) vaccine, mRNA-1273 0.05 MG/ML / SARS-CoV-2 (COVID-19) vaccine, mRNA-1273 OMICRON (BA.4/BA.5) 0.05 MG/ML Injectable Suspension |
| 779413 | SARS-CoV-2 (COVID-19) vaccine, mRNA-1273 0.1 MG/ML |
| 42631341 | SARS-CoV-2 (COVID-19) vaccine, mRNA-1273 0.1 MG/ML Injectable Suspension |
| 37003517 | SARS-CoV-2 (COVID-19) vaccine, mRNA-1273 0.2 MG/ML |
| 779676 | SARS-CoV-2 (COVID-19) vaccine, mRNA-1273 0.2 MG/ML [Spikevax] |
| 42796198 | SARS-CoV-2 (COVID-19) vaccine, mRNA-1273 0.2 MG/ML Injectable Suspension |
| 779679 | SARS-CoV-2 (COVID-19) vaccine, mRNA-1273 0.2 MG/ML Injectable Suspension [Spikevax] |
| 1525541 | SARS-CoV-2 (COVID-19) vaccine, mRNA-1273 OMICRON (BA.4/BA.5) |
| 742037 | SARS-CoV-2 (COVID-19) vaccine, mRNA-1273 OMICRON (BA.4/BA.5) 0.025 MG/ML |
| 1525542 | SARS-CoV-2 (COVID-19) vaccine, mRNA-1273 OMICRON (BA.4/BA.5) 0.05 MG/ML |
| 37003431 | Immunization administration by intramuscular injection of severe acute respiratory syndrome coronavirus 2 (SARS-CoV-2) (coronavirus disease [COVID-19]) vaccine, DNA, spike protein, adenovirus type 26 (Ad26) vector, preservative free, 5x1010 viral particles/0.5 mL dosage; booster dose |
| 742039 | SARS-CoV-2 (COVID-19) vaccine, mRNA-BNT162b2 0.0075 MG/ML |
| 36873496 | SARS-CoV-2 (COVID-19) vaccine, mRNA-BNT162b2 0.0075 MG/ML / SARS-CoV-2 (COVID-19) vaccine, mRNA-BNT162b2 OMICRON (BA.4/BA.5) 0.0075 MG/ML Injectable Suspension |
| 779947 | SARS-CoV-2 (COVID-19) vaccine, mRNA-BNT162b2 0.015 MG/ML |
| 1799557 | SARS-CoV-2 (COVID-19) vaccine, mRNA-BNT162b2 0.015 MG/ML Injectable Suspension |
| 742008 | SARS-CoV-2 (COVID-19) vaccine, mRNA-BNT162b2 0.025 MG/ML |
| 36873497 | SARS-CoV-2 (COVID-19) vaccine, mRNA-BNT162b2 0.025 MG/ML / SARS-CoV-2 (COVID-19) vaccine, mRNA-BNT162b2 OMICRON (BA.4/BA.5) 0.025 MG/ML Injectable Suspension |
| 702117 | SARS-CoV-2 (COVID-19) vaccine, mRNA-BNT162b2 0.05 MG/ML |
| 36873120 | SARS-CoV-2 (COVID-19) vaccine, mRNA-BNT162b2 0.05 MG/ML / SARS-CoV-2 (COVID-19) vaccine, mRNA-BNT162b2 OMICRON (BA.4/BA.5) 0.05 MG/ML Injectable Suspension |
| 1525545 | SARS-CoV-2 (COVID-19) vaccine, mRNA-BNT162b2 0.05 MG/ML / SARS-CoV-2 (COVID-19) vaccine, mRNA-BNT162b2 OMICRON (BA.4/BA.5) 0.05 MG/ML Injection |
| 1799562 | SARS-CoV-2 (COVID-19) vaccine, mRNA-BNT162b2 0.05 MG/ML Injectable Suspension |
| 37003433 | SARS-CoV-2 (COVID-19) vaccine, mRNA-BNT162b2 0.1 MG/ML |
| 1759203 | SARS-CoV-2 (COVID-19) vaccine, mRNA-BNT162b2 0.1 MG/ML [Comirnaty] |
| 42797616 | SARS-CoV-2 (COVID-19) vaccine, mRNA-BNT162b2 0.1 MG/ML Injectable Suspension |
| 36900252 | SARS-CoV-2 (COVID-19) vaccine, mRNA-BNT162b2 0.1 MG/ML Injectable Suspension [Comirnaty] |
| 1525509 | SARS-CoV-2 (COVID-19) vaccine, mRNA-BNT162b2 OMICRON (BA.4/BA.5) |
| 742040 | SARS-CoV-2 (COVID-19) vaccine, mRNA-BNT162b2 OMICRON (BA.4/BA.5) 0.0075 MG/ML |
| 1525544 | SARS-CoV-2 (COVID-19) vaccine, mRNA-BNT162b2 OMICRON (BA.4/BA.5) 0.05 MG/ML |
| 780154 | SARS-COV-2 (COVID-19) vaccine, subunit, recombinant spike protein Injectable Product |
| 780155 | SARS-COV-2 (COVID-19) vaccine, subunit, recombinant spike protein Injectable Suspension |
| 702679 | SARS-COV-2 (COVID-19) vaccine, subunit, recombinant spike protein-nanoparticle+Matrix-M1 Adjuvant, preservative free, 0.5mL dose |
| 739901 | SARS-COV-2 (COVID-19) vaccine, vector - Ad26 |
| 739903 | SARS-COV-2 (COVID-19) vaccine, vector - Ad26 100000000000 UNT/ML |
| 1226309 | SARS-COV-2 (COVID-19) vaccine, vector - Ad26 100000000000 UNT/ML Injectable Suspension |
| 766241 | Severeacuterespiratorysyndromecoronavirus2(SARS-CoV-2)(coronavirusdisease[COVID-19])vaccine,DNA,spikeprotein,adenovirustype26(Ad26)vector,preservativefree,5x1010viralparticles/0.5mLdosage,forintramuscularuse |
| 766240 | Severeacuterespiratorysyndromecoronavirus2(SARS-CoV-2)(coronavirusdisease[COVID-19])vaccine,DNA,spikeprotein,chimpanzeeadenovirusOxford1(ChAdOx1)vector,preservativefree,5x1010viralparticles/0.5mLdosage,forintramuscularuse |
| 759696 | Severeacuterespiratorysyndromecoronavirus2(SARS-CoV-2)(coronavirusdisease[COVID-19])vaccine,monovalent,preservativefree,5mcg/0.5mLdosage,adjuvantAS03emulsion,forintramuscularuse |
| 777323 | Severeacuterespiratorysyndromecoronavirus2(SARS-CoV-2)(coronavirusdisease[COVID-19])vaccine,mRNA-LNP,bivalentspikeprotein,preservativefree,10mcg/0.2mLdosage,diluentreconstituted,tris-sucroseformulation,forintramuscularuse |
| 733013 | Severeacuterespiratorysyndromecoronavirus2(SARS-CoV-2)(coronavirusdisease[COVID-19])vaccine,mRNA-LNP,bivalentspikeprotein,preservativefree,3mcg/0.2mLdosage,diluentreconstituted,tris-sucroseformulation,forintramuscularuse |
| 777320 | Severeacuterespiratorysyndromecoronavirus2(SARS-CoV-2)(coronavirusdisease[COVID-19])vaccine,mRNA-LNP,bivalentspikeprotein,preservativefree,30mcg/0.3mLdosage,tris-sucroseformulation,forintramuscularuse |
| 733014 | Severeacuterespiratorysyndromecoronavirus2(SARS-CoV-2)(coronavirusdisease[COVID-19])vaccine,mRNA-LNP,spikeprotein,bivalent,preservativefree,10mcg/0.2mLdosage,forintramuscularuse |
| 777322 | Severeacuterespiratorysyndromecoronavirus2(SARS-CoV-2)(coronavirusdisease[COVID-19])vaccine,mRNA-LNP,spikeprotein,bivalent,preservativefree,25mcg/0.25mLdosage,forintramuscularuse |
| 777321 | Severeacuterespiratorysyndromecoronavirus2(SARS-CoV-2)(coronavirusdisease[COVID-19])vaccine,mRNA-LNP,spikeprotein,bivalent,preservativefree,50mcg/0.5mLdosage,forintramuscularuse |
| 759738 | Severeacuterespiratorysyndromecoronavirus2(SARS-CoV-2)(coronavirusdisease[COVID-19])vaccine,mRNA-LNP,spikeprotein,preservativefree,10mcg/0.2mLdosage,diluentreconstituted,tris-sucroseformulation,forintramuscularuse |
| 766239 | Severeacuterespiratorysyndromecoronavirus2(SARS-CoV-2)(coronavirusdisease[COVID-19])vaccine,mRNA-LNP,spikeprotein,preservativefree,100mcg/0.5mLdosage,forintramuscularuse |
| 777319 | Severeacuterespiratorysyndromecoronavirus2(SARS-CoV-2)(coronavirusdisease[COVID-19])vaccine,mRNA-LNP,spikeprotein,preservativefree,25mcg/0.25mLdosage,forintramuscularuse |
| 759694 | Severeacuterespiratorysyndromecoronavirus2(SARS-CoV-2)(coronavirusdisease[COVID-19])vaccine,mRNA-LNP,spikeprotein,preservativefree,3mcg/0.2mLdosage,diluentreconstituted,tris-sucroseformulation,forintramuscularuse |
| 766238 | Severeacuterespiratorysyndromecoronavirus2(SARS-CoV-2)(coronavirusdisease[COVID-19])vaccine,mRNA-LNP,spikeprotein,preservativefree,30mcg/0.3mLdosage,diluentreconstituted,forintramuscularuse |
| 759736 | Severeacuterespiratorysyndromecoronavirus2(SARS-CoV-2)(coronavirusdisease[COVID-19])vaccine,mRNA-LNP,spikeprotein,preservativefree,30mcg/0.3mLdosage,tris-sucroseformulation,forintramuscularuse |
| 759737 | Severeacuterespiratorysyndromecoronavirus2(SARS-CoV-2)(coronavirusdisease[COVID-19])vaccine,mRNA-LNP,spikeprotein,preservativefree,50mcg/0.25mLdosage,forintramuscularuse |
| 759695 | Severeacuterespiratorysyndromecoronavirus2(SARS-CoV-2)(coronavirusdisease[COVID-19])vaccine,mRNA-LNP,spikeprotein,preservativefree,50mcg/0.5mLdosage,forintramuscularuse |
| 759735 | Severeacuterespiratorysyndromecoronavirus2(SARS-CoV-2)(coronavirusdisease[COVID-19])vaccine,recombinantspikeproteinnanoparticle,saponin-basedadjuvant,preservativefree,5mcg/0.5mLdosage,forintramuscularuse |
| 702866 | SARS-COV-2(COVID-19)vaccine,vectornon-replicating,recombinantspikeprotein-Ad26,preservativefree,0.5mL |

| **Supplementary Table 1.b: COVID-19 Infection Diagnostic Concept IDs** | |
| --- | --- |
| **Concept IDs** | **Concept name** |
| 3661405 | Acute bronchitis caused by SARS-CoV-2 |
| 3655976 | Acute hypoxemic respiratory failure due to disease caused by severe acute respiratory syndrome coronavirus 2 |
| 3661748 | Acute kidney injury due to disease caused by severe acute respiratory syndrome coronavirus 2 |
| 756044 | Acute respiratory distress syndrome (ARDS) caused by COVID-19 |
| 3661406 | Acute respiratory distress syndrome due to disease caused by severe acute respiratory syndrome coronavirus 2 |
| 756061 | Asymptomatic COVID-19 |
| 3662381 | Asymptomatic SARS-CoV-2 |
| 756031 | Bronchitis caused by COVID-19 |
| 3656667 | Cardiomyopathy due to disease caused by severe acute respiratory syndrome coronavirus 2 |
| 3656668 | Conjunctivitis due to disease caused by severe acute respiratory syndrome coronavirus 2 |
| 37311061 | COVID-19 |
| 37310269 | COVID-19 |
| 703441 | COVID-19 confirmed by laboratory test |
| 3656669 | Dyspnea caused by severe acute respiratory syndrome coronavirus 2 |
| 702953 | Emergency use of U07.1 \| COVID-19 |
| 45756093 | Emergency use of U07.1 \| COVID-19, virus identified |
| 37310284 | Encephalopathy due to disease caused by severe acute respiratory syndrome coronavirus 2 |
| 3661885 | Fever caused by severe acute respiratory syndrome coronavirus 2 |
| 37310283 | Gastroenteritis caused by SARS-CoV-2 (severe acute respiratory syndrome coronavirus 2) |
| 756081 | Infection of lower respiratory tract caused by COVID-19 |
| 37310286 | Infection of upper respiratory tract caused by severe acute respiratory syndrome coronavirus 2 |
| 3663281 | Lower respiratory infection caused by SARS-CoV-2 |
| 3661631 | Lymphocytopenia due to severe acute respiratory syndrome coronavirus 2 |
| 37310287 | Myocarditis due to disease caused by severe acute respiratory syndrome coronavirus 2 |
| 37310254 | Otitis media due to disease caused by severe acute respiratory syndrome coronavirus 2 |
| 713860 | Personal history of COVID-19 |
| 3661408 | Pneumonia caused by SARS-CoV-2 |
| 713857 | Pneumonia due to coronavirus disease 2019 |
| 766502 | Post COVID-19 condition |
| 710708 | Post COVID-19 condition |
| 766503 | Post COVID-19 condition, unspecified |
| 705076 | Post-acute COVID-19 |
| 756039 | Respiratory infection caused by COVID-19 |
| 3655977 | Rhabdomyolysis due to disease caused by severe acute respiratory syndrome coronavirus 2 |
| 3655975 | Sepsis due to disease caused by severe acute respiratory syndrome coronavirus 2 |
| 3661632 | Thrombocytopenia due to severe acute respiratory syndrome coronavirus 2 |

| **Supplementary Table 1.c: COVID-19 Infection Laboratory Test Concept IDs** | |
| --- | --- |
| **Concept IDs** | **Concept name** |
| 756055 | Measurement of Severe acute respiratory syndrome coronavirus 2 (SARS-CoV-2) |
| 40763330 | SARS coronavirus RNA [Presence] in Isolate by NAA with probe detection |
| 3031852 | SARS coronavirus RNA [Presence] in Specimen by NAA with probe detection |
| 586515 | SARS-CoV-2 (COVID-19) Ab [Presence] in Serum or Plasma by Immunoassay |
| 586522 | SARS-CoV-2 (COVID-19) Ab [Units/volume] in Serum or Plasma by Immunoassay |
| 706179 | SARS-CoV-2 (COVID-19) Ab panel - Serum or Plasma by Immunoassay |
| 706176 | SARS-CoV-2 (COVID-19) Ab panel - Serum, Plasma or Blood by Rapid immunoassay |
| 36032419 | SARS-CoV-2 (COVID-19) Ag [Presence] in Upper respiratory specimen by Immunoassay |
| 36033641 | SARS-CoV-2 (COVID-19) Ag [Presence] in Upper respiratory specimen by Rapid immunoassay |
| 723473 | SARS-CoV-2 (COVID-19) IgA Ab [Presence] in Serum or Plasma by Immunoassay |
| 586527 | SARS-CoV-2 (COVID-19) IgG Ab [Presence] in DBS by Immunoassay |
| 706181 | SARS-CoV-2 (COVID-19) IgG Ab [Presence] in Serum, Plasma or Blood by Rapid immunoassay |
| 706177 | SARS-CoV-2 (COVID-19) IgG Ab [Units/volume] in Serum or Plasma by Immunoassay |
| 723479 | SARS-CoV-2 (COVID-19) IgG+IgM Ab [Presence] in Serum or Plasma by Immunoassay |
| 723475 | SARS-CoV-2 (COVID-19) IgM Ab [Presence] in Serum or Plasma by Immunoassay |
| 706180 | SARS-CoV-2 (COVID-19) IgM Ab [Presence] in Serum, Plasma or Blood by Rapid immunoassay |
| 706178 | SARS-CoV-2 (COVID-19) IgM Ab [Units/volume] in Serum or Plasma by Immunoassay |
| 706157 | SARS-CoV-2 (COVID-19) N gene [Cycle Threshold #] in Specimen by Nucleic acid amplification using CDC primer-probe set N1 |
| 706155 | SARS-CoV-2 (COVID-19) N gene [Cycle Threshold #] in Specimen by Nucleic acid amplification using CDC primer-probe set N2 |
| 715272 | SARS-CoV-2 (COVID-19) N gene [Presence] in Nasopharynx by NAA with probe detection |
| 36033644 | SARS-CoV-2 (COVID-19) N gene [Presence] in Nose by NAA with non-probe detection |
| 757678 | SARS-CoV-2 (COVID-19) N gene [Presence] in Nose by NAA with probe detection |
| 36661378 | SARS-CoV-2 (COVID-19) N gene [Presence] in Saliva (oral fluid) by NAA with probe detection |
| 36032258 | SARS-CoV-2 (COVID-19) N gene [Presence] in Saliva (oral fluid) by Nucleic acid amplification using CDC primer-probe set N1 |
| 706175 | SARS-CoV-2 (COVID-19) N gene [Presence] in Specimen by NAA with probe detection |
| 706156 | SARS-CoV-2 (COVID-19) N gene [Presence] in Specimen by Nucleic acid amplification using CDC primer-probe set N1 |
| 706154 | SARS-CoV-2 (COVID-19) N gene [Presence] in Specimen by Nucleic acid amplification using CDC primer-probe set N2 |
| 36033640 | SARS-CoV-2 (COVID-19) ORF1ab region [Units/volume] (viral load) in Upper respiratory specimen by NAA with probe detection |
| 715262 | SARS-CoV-2 (COVID-19) RNA [Log #/volume] (viral load) in Specimen by NAA with probe detection |
| 723476 | SARS-CoV-2 (COVID-19) RNA [Presence] in Nasopharynx by NAA with non-probe detection |
| 586526 | SARS-CoV-2 (COVID-19) RNA [Presence] in Nasopharynx by NAA with probe detection |
| 757677 | SARS-CoV-2 (COVID-19) RNA [Presence] in Nose by NAA with probe detection |
| 715260 | SARS-CoV-2 (COVID-19) RNA [Presence] in Saliva (oral fluid) by NAA with probe detection |
| 706170 | SARS-CoV-2 (COVID-19) RNA [Presence] in Specimen by NAA with probe detection |
| 706169 | SARS-CoV-2 (COVID-19) RNA panel - Specimen by NAA with probe detection |
| 723470 | SARS-CoV-2 (COVID-19) RdRp gene [Cycle Threshold #] in Specimen by NAA with probe detection |
| 706173 | SARS-CoV-2 (COVID-19) RdRp gene [Presence] in Specimen by NAA with probe detection |
| 586516 | SARS-CoV-2 (COVID-19) [Presence] in Specimen by Organism specific culture |
| 757680 | SARS-CoV-2 (COVID-19) neutralizing antibody [Presence] in Serum by pVNT |
| 757679 | SARS-CoV-2 (COVID-19) neutralizing antibody [Titer] in Serum by pVNT |
| 36031944 | SARS-CoV-2 (COVID-19) specific TCRB gene rearrangements [Presence] in Blood by Sequencing |
| 36033667 | SARS-CoV-2 (COVID-19) variant [Type] in Specimen by Sequencing |
| 706174 | SARS-related coronavirus E gene [Presence] in Specimen by NAA with probe detection |
| 706171 | SARS-related coronavirus N gene [Presence] in Specimen by Nucleic acid amplification using CDC primer-probe set N3 |
| 723472 | SARS-related coronavirus RNA [Presence] in Specimen by NAA with probe detection |

| **Supplementary Table 1.d: Well-Care Visit Concept IDs** | |
| --- | --- |
| **Concept IDs** | **Concept name** |
| 4301122 | History and physical examination, annual for health maintenance |
| 4094041 | Well woman health examination |
| 4194672 | Specialized medical examination |
| 4061660 | Preventive procedure |
| 4104747 | Physical examination, complete |
| 4183687 | History and physical examination, administrative |
| 4145333 | Adult health examination |
| 4094039 | Well man health examination |
| 4065061 | Individual health examination |
| 4036803 | General examination of patient |
| 32693 | Health examination |
| 4240345 | Physical examination |
| 4065062 | Health examination of sub-group |
| 4062858 | Individual general health examination |

| **Supplementary Table 1.e: GBS Diagnostic Concept IDs** | |
| --- | --- |
| **Concept IDs** | **Concept name** |
| 4164770 | Guillain-Barré syndrome |
| 37396086 | Acute motor axonal neuropathy |
| 4070552 | Fisher’s syndrome |
| 37396786 | Acute inflammatory demyelinating polyradiculoneuropathy |
| 37204369 | Pharyngeal-cervical-brachial variant of Guillain-Barré syndrome |
| 35624130 | Paraparetic variant of Guillain-Barré syndrome |
| 35623634 | Acute sensory ataxic neuropathy |
| 37396785 | [Acute motor sensory axonal neuropathy](https://athena.ohdsi.org/search-terms/terms/37396785) |
| 40322804 | [Acute infective polyneuritis (& [Guillain-Barre syndrome])](https://athena.ohdsi.org/search-terms/terms/40322804) |
| 3407887 | [Acute inflammatory demyelinating polyradiculoneuropathy](https://athena.ohdsi.org/search-terms/terms/3407887) |
| 1411961 | [Fisher syndrome (machine translation)](https://athena.ohdsi.org/search-terms/terms/1411961) |
| 1411962 | [Guillain - Barre syndrome axonal (machine translation)](https://athena.ohdsi.org/search-terms/terms/1411962) |
| 1411963 | [Guillain - Barre syndrome, demyelinating (machine translation)](https://athena.ohdsi.org/search-terms/terms/1411963) |
| 45615949 | [Guillain-Barre Syndrome](https://athena.ohdsi.org/search-terms/terms/45615949) |
| 35207452 | [Guillain-Barre Syndrome](https://athena.ohdsi.org/search-terms/terms/45615949) |
| 42487250 | [Guillain-Barre Syndrome](https://athena.ohdsi.org/search-terms/terms/45615949) |
| 45463400 | [Guillain-Barre Syndrome](https://athena.ohdsi.org/search-terms/terms/45615949) |
| 1411959 | [Guillain-Barre Syndrome](https://athena.ohdsi.org/search-terms/terms/45615949) |
| 1411960 | [Guillain-Barre Syndrome](https://athena.ohdsi.org/search-terms/terms/45615949) |
| 37083560 | [Guillain-Barre Syndrome](https://athena.ohdsi.org/search-terms/terms/45615949) |
| 37610725 | [Guillain-Barre Syndrome](https://athena.ohdsi.org/search-terms/terms/45615949) |
| 45562102 | [Guillain-Barre Syndrome](https://athena.ohdsi.org/search-terms/terms/45615949) |

**Supplementary Figure 1: Forest Plot of Incidence Risk Ratios for GBS After COVID-19 Vaccination or Infection During 30-Day Follow-up**

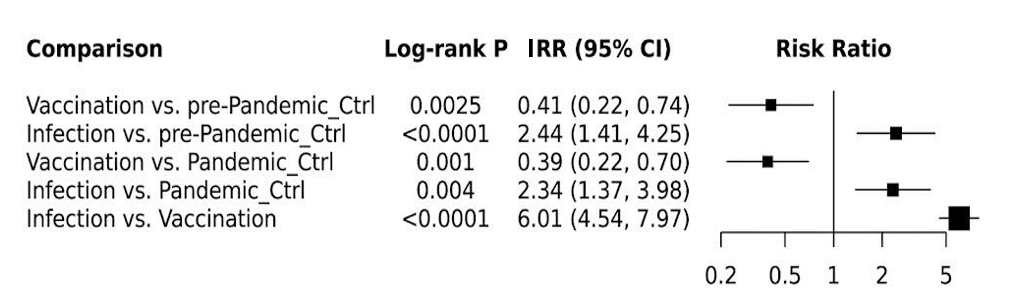

Pairwise comparisons of risk ratios (RRs) for GBS following COVID-19 vaccination or SARS-CoV-2 infection relative to pre-pandemic and pandemic controls. Vaccination was associated with a reduced risk of GBS, whereas COVID-19 infection was associated with an increased risk. Vaccination was associated with a significantly lower risk of GBS compared to pre-pandemic controls, Pandemic controls, and Infection. Infection was associated with more than twice the GBS risk compared to Pre-Pandemic and Pandemic controls. Squares represent RR estimates and horizontal lines indicate 95% CIs. The vertical reference line represents RR = 1. Abbreviations: IRR, incidence risk ratio; CI, confidence interval; GBS, Guillain-Barré Syndrome**.**

**Supplementary Figure 2: Association Between COVID-19 Vaccination and GBS Incidence Across Post-Vaccination Follow-Up Periods Using a Multivariable-Adjusted Cox Model**

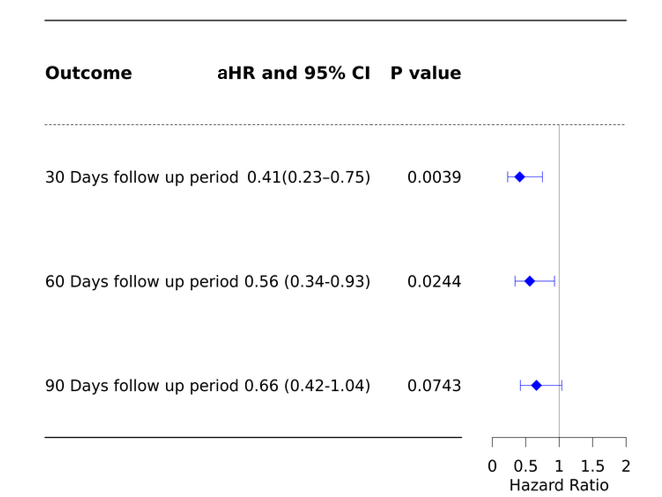

Multivariable-adjusted Cox proportional hazards model was used to assess the association between COVID-19 vaccination and GBS risk. The primary analysis evaluated outcomes over a 30-day follow-up period, with extended analyses at 60 and 90 days post-index date. Adults were categorized into two groups based on initial COVID-19 vaccination exposure from December 2020 to November 2024. The vaccination group included individuals receiving their first COVID-19 vaccine dose before any infection, while the control group consisted of individuals with a well-care visit before any COVID-19 vaccination or infection. Each row represents an independent outcome. Abbreviations: aHR, adjusted hazard ratio; CI, confidence interval; GBS, Guillain-Barré Syndrome.

**Supplementary Figure 3: Risk of GBS Within 30 Days After COVID-19 Vaccination Stratified by Doses or Co-administered Vaccines**
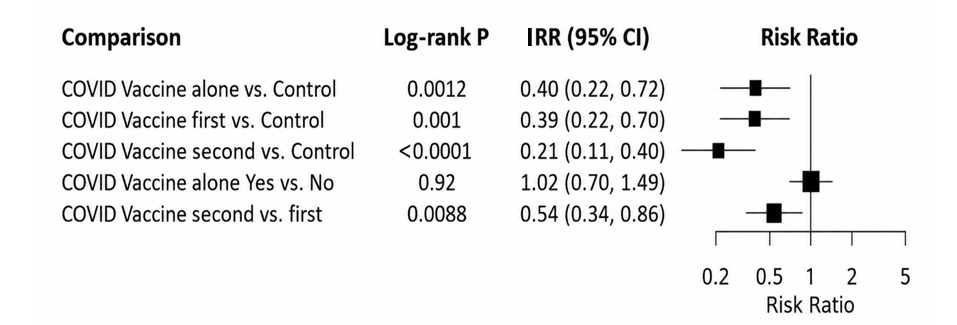

Forest plot showing pairwise comparisons of risk ratios (RRs) for GBS within 30 days after COVID-19 vaccination. Analyses were stratified by vaccine dose number and by exclusion of co-administered vaccines. Second-dose vaccination was associated with a significantly lower risk of GBS compared with controls and first-dose vaccination, while exclusion of co-administered vaccines did not significantly alter risk estimates. Squares represent RR estimates and horizontal lines indicate 95% CIs. The vertical reference line represents RR = 1. Abbreviations: IRR, incidence risk ratio; CI, confidence interval; GBS, Guillain-Barré Syndrome.

**Supplementary Figure 4: Comparison of GBS Risk Following COVID-19 mRNA and Adenoviral Vector Vaccination During 30-Day Follow-up Period**
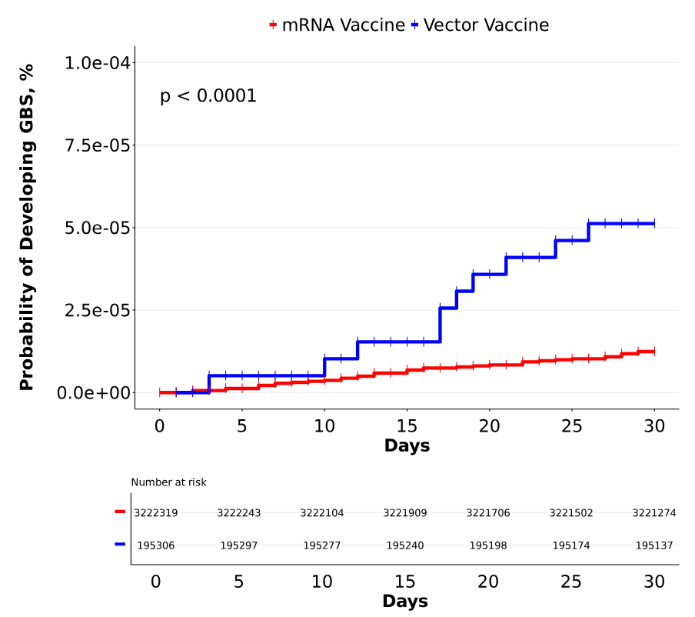

Cumulative probability curves illustrate the likelihood of developing GBS over a 30-day follow-up period after COVID-19 Vaccination. The red and blue curves depict individuals who received COVID-19 mRNA or adenoviral vector vaccines respectively. The x-axis represents days from index date, and the y-axis indicates the probability of developing GBS as a percentage. Statistical significance was assessed using a Log-rank test. Abbreviation: GBS, Guillain-Barré Syndrome.

**Supplementary Figure 5: Association Between COVID-19 Vaccine Manufacturers and Risk of GBS Within 30 Days of Vaccination**

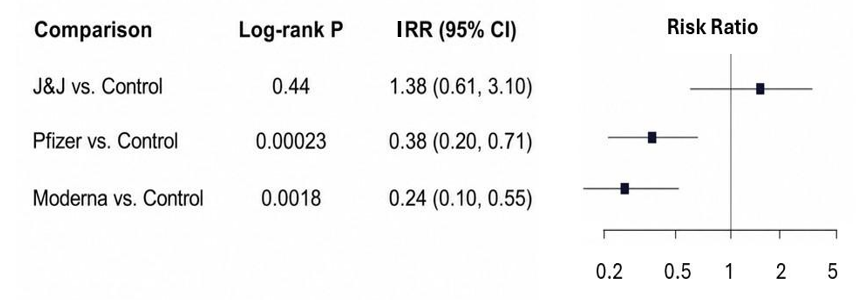

Forest plot displaying the risk ratios (RR) and 95% CIs for GBS diagnosis following COVID-19 infection among individuals previously vaccinated with J&J, Pfizer, or Moderna vaccines compared to unvaccinated controls. Log-rank *P*-values indicate the statistical significance of each comparison. Squares represent RR estimates and horizontal lines indicate 95% CIs. The vertical reference line represents RR = 1. Abbreviations: IRR, incidence risk ratio; CI, confidence interval; GBS, Guillain-Barré Syndrome

**Supplementary Figure 6: Association of GBS Risk Within 30 Days Following COVID-19 Vaccination After Excluding Other Vaccine Coadministration**

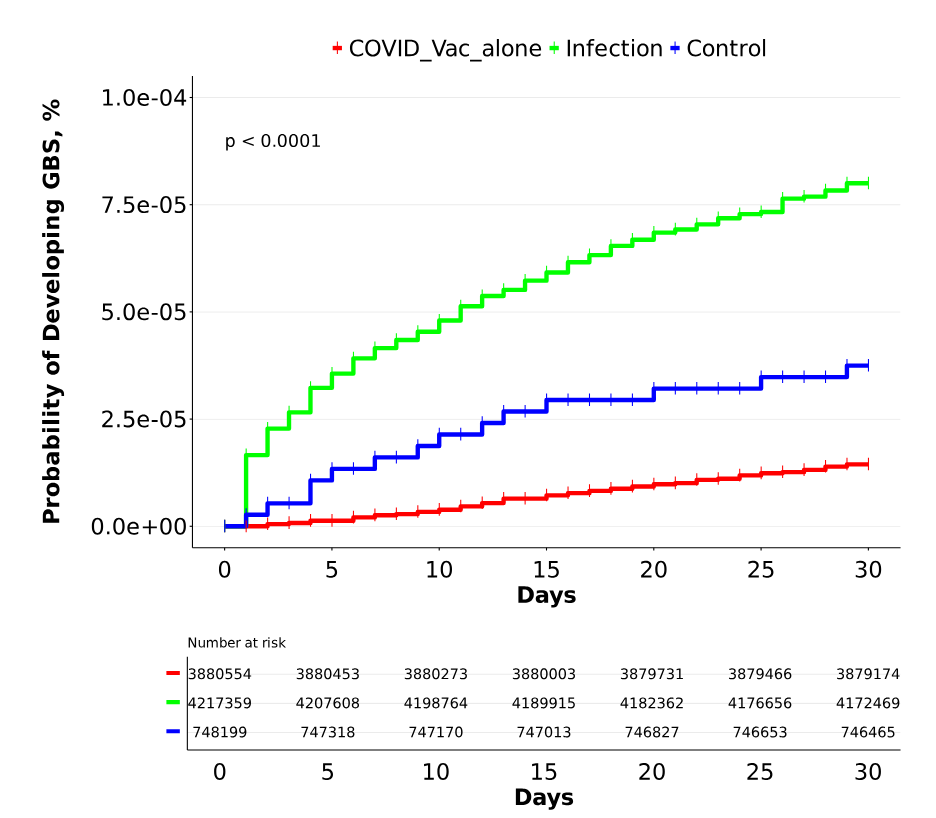

Cumulative probability curves illustrate the likelihood of developing GBS over a 30-day follow-up period. The green curve represents the COVID-19 infection group, while the red and blue curves depict individuals who received COVID-19 vaccines alone or unvaccinated control respectively. The x-axis represents days from index date, and the y-axis indicates the probability of developing GBS as a percentage. Statistical significance was assessed using a Log-rank test. Abbreviations: GBS, Guillain-Barré Syndrome.

**Supplementary Figure 7: Risk of GBS Following COVID-19 Infection Stratified by Pre-Infection Vaccination-to-Infection Time Interval**

a

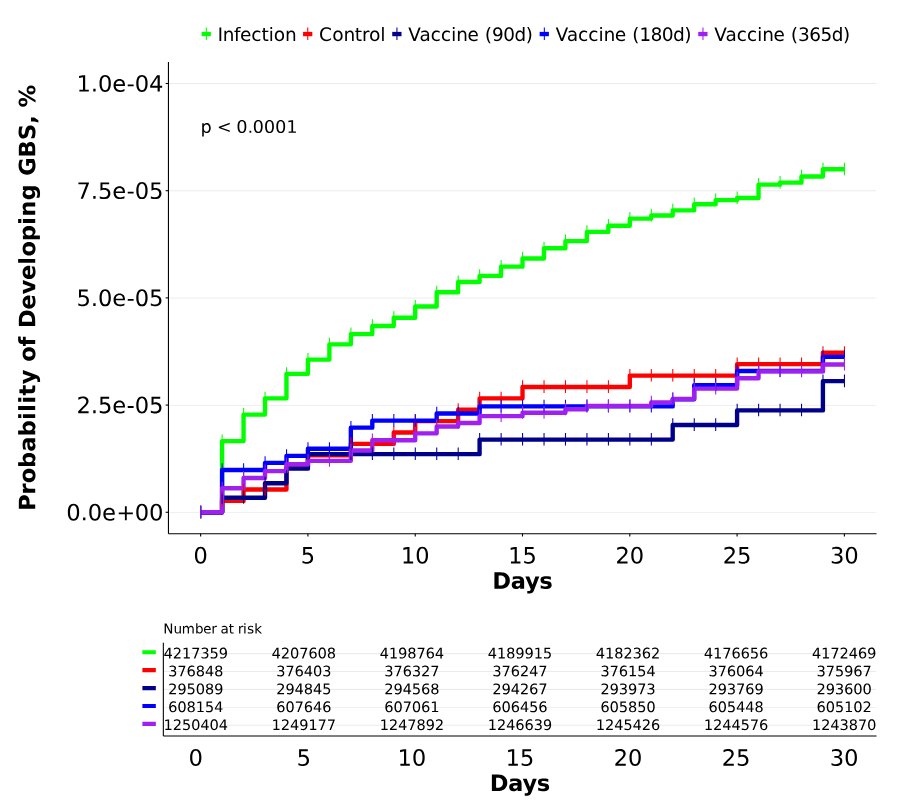

b

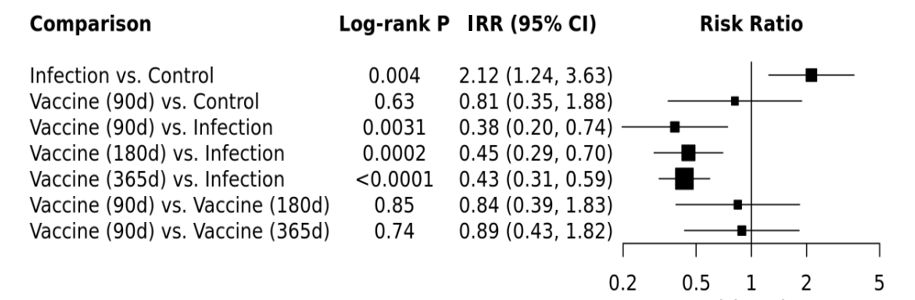

Risk of GBS following COVID-19 infection, stratified by prior infection vaccination-to-infection interval. (a) Kaplan–Meier curves depict the 30-day cumulative incidence of GBS. The groups include unvaccinated infection-only individuals (green curve), unexposed controls (red curve), and infected individuals stratified by time since vaccination: 90 days (dark blue curve), 180 days (blue curve), and 365 days (purple curve). The x-axis represents days from the index date, and the y-axis shows the cumulative probability of developing GBS in percentage. Differences between curves were evaluated using the Log-rank test. (b) Forest plot illustrates the risk ratios for GBS. The model compares the risk of GBS among infected individuals who were vaccinated at different intervals (90, 180, and 365 days prior COVID-19 infection) with that of the unvaccinated infection group. Pairwise comparisons among the vaccinated groups were presented. All prior infection vaccinated groups demonstrated a significantly lower hazard of GBS compared with the unvaccinated infection group. No significant differences were observed among the prior vaccinated groups, suggesting consistent protective effects across the examined time intervals. Squares represent RR estimates and horizontal lines indicate 95% CIs. The vertical reference line represents RR = 1. Abbreviations: HR, hazard ratio; CI, confidence interval. Abbreviations: IRR, incidence risk ratio; CI, confidence interval; GBS, Guillain-Barré Syndrome.

**Supplementary Figure 8: Association Between COVID-19 Infection and GBS Risk Evaluated Using a Multivariable-Adjusted Cox Proportional Hazards Model**

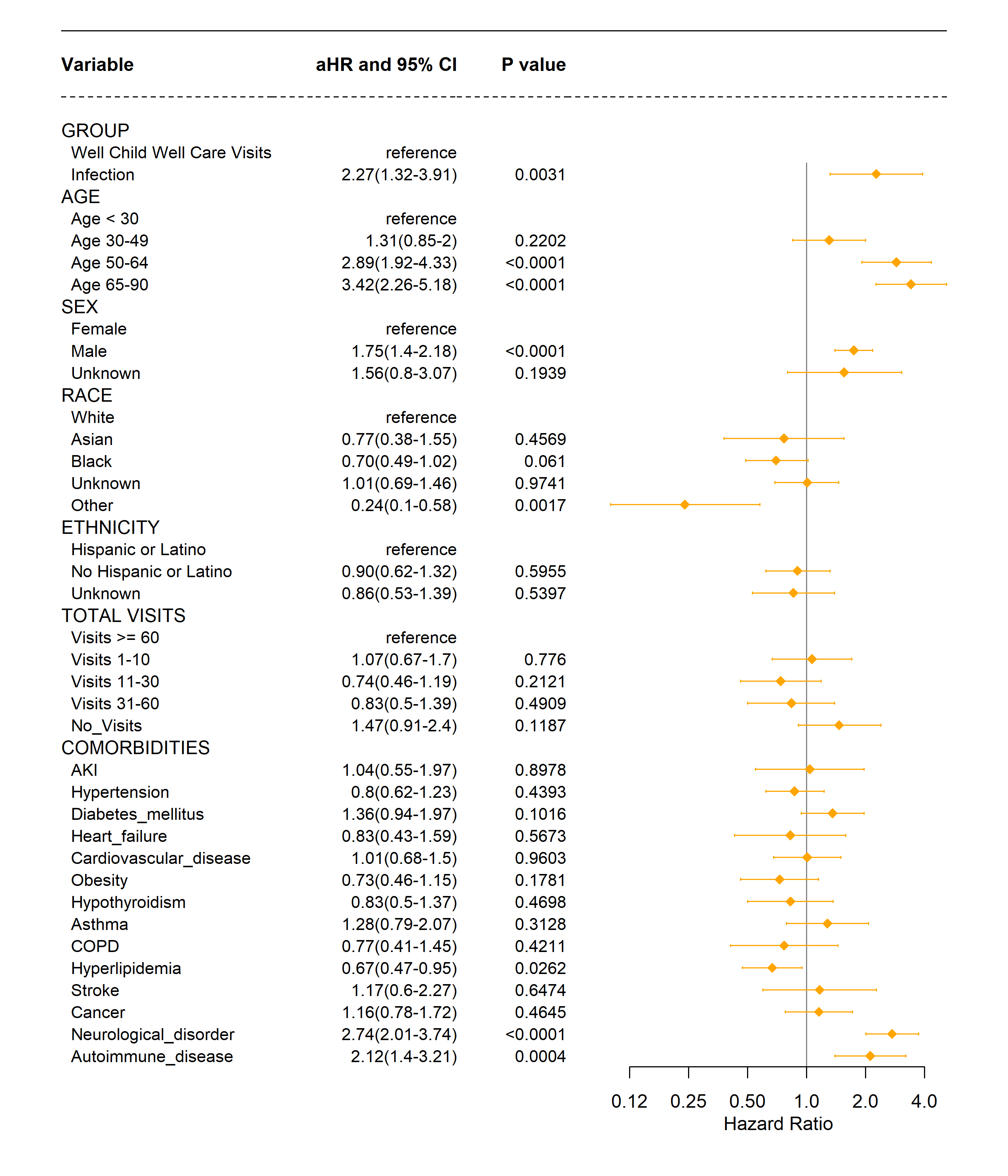

Multivariable Cox proportional hazards model adjusted for age, gender, race, total visits, and comorbidities, was used to compare the COVID-19 infection and control groups. Adults were categorized into two groups based on their initial COVID-19 exposure between December 2020 and November 2024. Each table row represents an independent outcome, with the primary outcome evaluated over a 30-day follow-up period. Abbreviations: aHR, adjusted hazard ratio; CI, confidence interval.
